# Positive Airway Pressure Therapy Predicts Lower Mortality and Major Adverse Cardiovascular Events Incidence in Medicare Beneficiaries with Obstructive Sleep Apnea

**DOI:** 10.1101/2023.07.26.23293156

**Authors:** Diego R Mazzotti, Lemuel R. Waitman, Jennifer Miller, Krishna M. Sundar, Nancy H. Stewart, David Gozal, Xing Song, Greater Plains Collaborative

## Abstract

**Background:** Obesity is associated with obstructive sleep apnea (OSA) and cardiovascular risk. Positive airway pressure (PAP) is the first line treatment for OSA, but evidence on its beneficial effect on major adverse cardiovascular events (MACE) prevention is limited. Using claims data, the effects of PAP on mortality and incidence of MACE among Medicare beneficiaries with OSA were examined.

**Methods:** A cohort of Medicare beneficiaries with ≥2 distinct OSA claims was defined from multi-state, state-wide, multi-year (2011-2020) Medicare fee-for-service claims data. Evidence of PAP initiation and utilization was based on PAP claims after OSA diagnosis. MACE was defined as a composite of myocardial infarction, heart failure, stroke, or coronary revascularization. Doubly robust Cox proportional hazards models with inverse probability of treatment weights estimated treatment effects controlling for sociodemographic and clinical factors.

**Results:** Among 888,835 beneficiaries with OSA (median age 73 years; 43.9% women; median follow-up 1,141 days), those with evidence of PAP initiation (32.6%) had significantly lower all-cause mortality (HR [95%CI]: 0.53 [0.52-0.54]) and MACE incidence risk (0.90 [0.89-0.91]). Higher quartiles of annual PAP claims were progressively associated with lower mortality (Q2: 0.84 [0.81-0.87], Q3: 0.76 [0.74-0.79], Q4: 0.74 [0.72-0.77]) and MACE incidence risk (Q2: 0.92 [0.89-0.95], Q3: 0.89 [0.86-0.91], Q4: 0.87 [0.85-0.90]).

**Conclusion:** PAP utilization was associated with lower all-cause mortality and MACE incidence among Medicare beneficiaries with OSA. Results might inform trials assessing the importance of OSA therapy towards minimizing cardiovascular risk and mortality in older adults.

## Introduction

Obesity is a chronic condition with significant public health burden^1^, affecting 40% of adults in the U.S.^2^ and accounting for millions of cardiovascular (CV) disease deaths every year^3^. There is greater attention about the importance of adequate sleep habits^4^ and its relation to cardiometabolic risk^5,6^. Obstructive sleep apnea (OSA) is a highly prevalent disorder contributing to these associations, with population rates of 9%-37% in men and 4-50% in women^7^, affecting nearly 1 billion people worldwide^8^. OSA becomes more prevalent with age^9,10^ and obesity^11^, and is associated with coronary heart disease, stroke, and CV mortality^12,13^. The CV risk due to OSA is greater among those with excessive sleepiness^14,15^, worse nocturnal hypoxemia^16^, and differential heart rate responses to respiratory events^17^. Thus, there is increasing attention on therapies that may modify CV risk for primary and secondary prevention^5,6,18^ through targeting sleep disordered breathing^19^.

Positive airway pressure (PAP) is the first line of therapy for moderate-severe OSA. Despite epidemiological studies strongly suggesting that OSA is a modifiable CV risk factor^20–26^, a meta-analysis of randomized controlled trials (RCTs) failed to demonstrate that PAP prevents CV outcomes^27^. Studies have shown that patient selection and treatment adherence may explain some of the negative results^28,29^. These challenges are intensified by the lack of large, equitable, and multi-centric pragmatic studies assessing the effectiveness of PAP on improving CV risk. Such studies are expensive and time-consuming, leading to recruitment and retention challenges, and delayed policy changes^30,31^. Whether long-term PAP therapy prevents CV disease in a clinical population is a critical question that remains unanswered.

Insurance claims data allow the design of observational studies that complement RCTs, by adopting causal inference methods^29,32,33^. When robustly applied, they have the potential to characterize the effect of OSA therapies in representative clinical settings^34^. A study on a French nationwide claims database found that continuous PAP (CPAP) therapy termination was associated with all-cause mortality and incidence of heart failure^35^. In the U.S., Centers for Medicare & Medicaid Services (CMS) beneficiaries represent a large population of older adults with access to healthcare coverage. Studies exploring a 5% sample of randomly selected beneficiaries found that those with OSA had higher healthcare utilization when compared to matched controls^36^. Moreover, those adherent to CPAP based on Durable Medical Equipment (DME) claims were found to have reduced risk of stroke^37^ and healthcare expenses among those with pre-existing CV diseases^38^. However, a major limitation is the reliance only on a sample of beneficiaries, which might represent challenges characterizing the effects of PAP on the incidence of individual CV diseases with accuracy. State-wide Medicare claims, particularly within underserved states in the U.S., might reflect a broader representation of individuals with OSA and provide more generalizable effect estimates.

In this study, we aimed to determine the effect of PAP utilization on all-cause mortality and incidence of MACE and its components among Medicare beneficiaries in the Central U.S. By leveraging a robust causal inference design, we determined the average treatment effect of PAP initiation on the risk of incident MACE and mortality. Next, we established clinically relevant cut-offs of PAP utilization based on DME claims over the first year after PAP initiation to determine PAP utilization exposure groups. We demonstrate that these groups are differentially associated with incident MACE and mortality among older adults. Finally, we provide effect estimates of PAP exposure groups stratified by relevant sociodemographic and clinical factors.

## Methods

Additional details are presented in the ***Supplemental materials***.

### Study cohort

Medicare beneficiaries (>65 years) enrolled to part A and B, and ≥2 distinct OSA claims were collected from multi-state, state-wide, multi-year (2011-2020) Medicare fee-for-service claims data. State-level Medicare claims data were originally obtained as part of the Greater Plains Collaborative Reusable Observable Unified Study Environment (GROUSE)^39^, with a catchment area across 11 states in the Central U.S. The study protocol has been approved by Institutional Review Boards at each institution. The study cohort was defined based on a validated EHR algorithm to identify participants with OSA^40^ (see **Supplemental Table 1**). We further required >1 year enrolment with Medicare before their first OSA claim, to better capture newly diagnosed OSA and complete PAP utilization history. For the analysis of incident MACE, we further excluded beneficiaries with history of MACE prior to their OSA diagnosis date.

### Study design

This is an observational, retrospective analysis of state-wide Medicare claims data. We proposed two complementary causal inference designs to estimate the average treatment effect (ATE) of PAP initiation (**Figure 1**) and PAP utilization exposure groups (**Figure 2**). For the PAP initiation analyses, prescription time-distribution matching^41^ was used to identify the “time zero” (exposure assignment, covariate determination, and start of follow-up) for the group that did not initiate PAP (*pseudo landmark date*, **Figure 1**), matched on the distribution of the time from diagnosis and start of PAP therapy in the group that initiated PAP (*landmark T*, **Figure 1**). For the analysis of PAP utilization exposure groups, we included beneficiaries that have been enrolled and have not experienced events at the 1-year mark since their PAP initiation date. Exposure groups were defined based on the distribution of claim counts during this first year of PAP utilization and “time zero” was the date of the first anniversary of PAP initiation (**Figure 2**).

**Figure 1.**
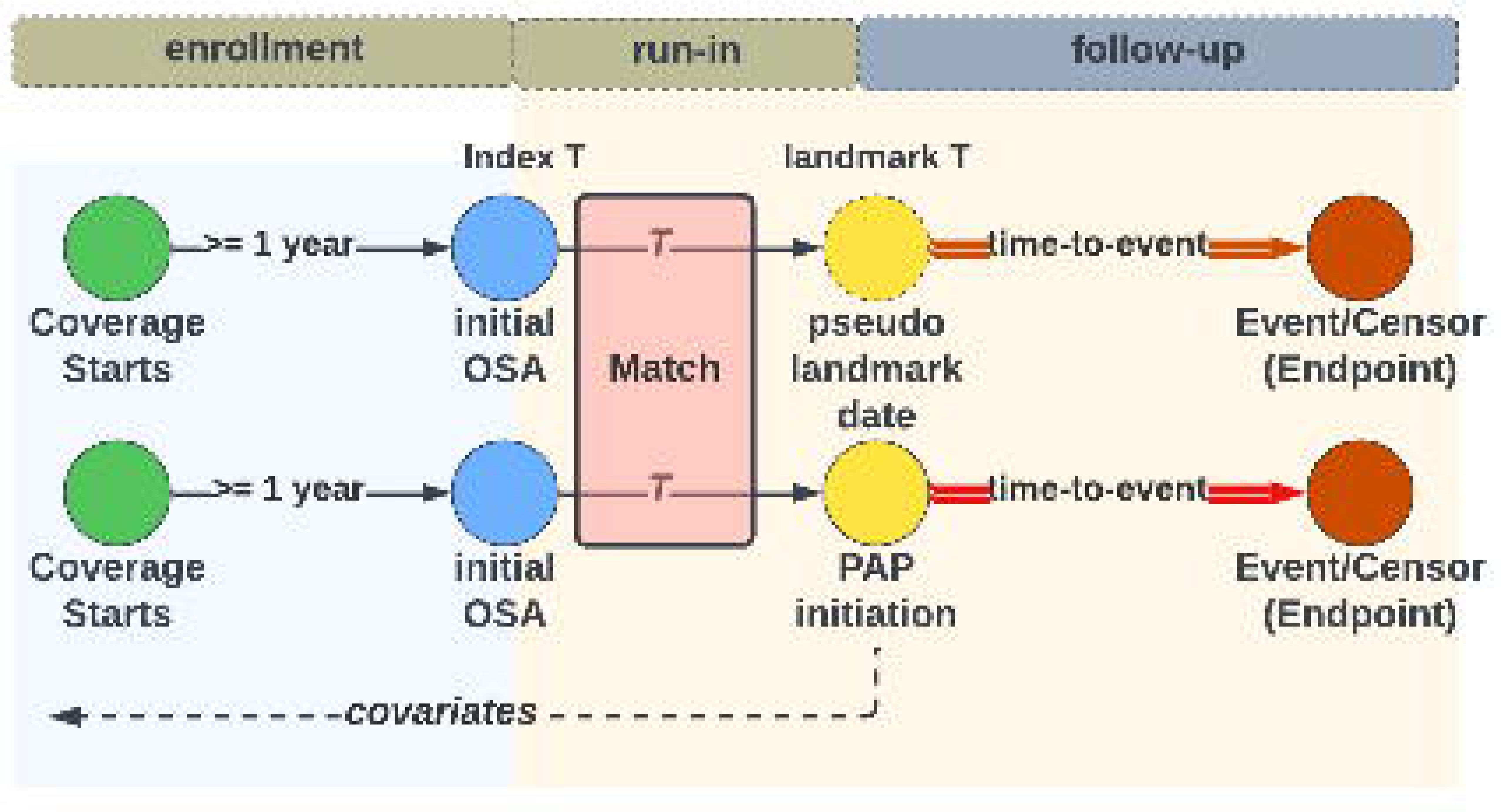
Overview of the PAP initiation analysis study design.

**Figure 2.**
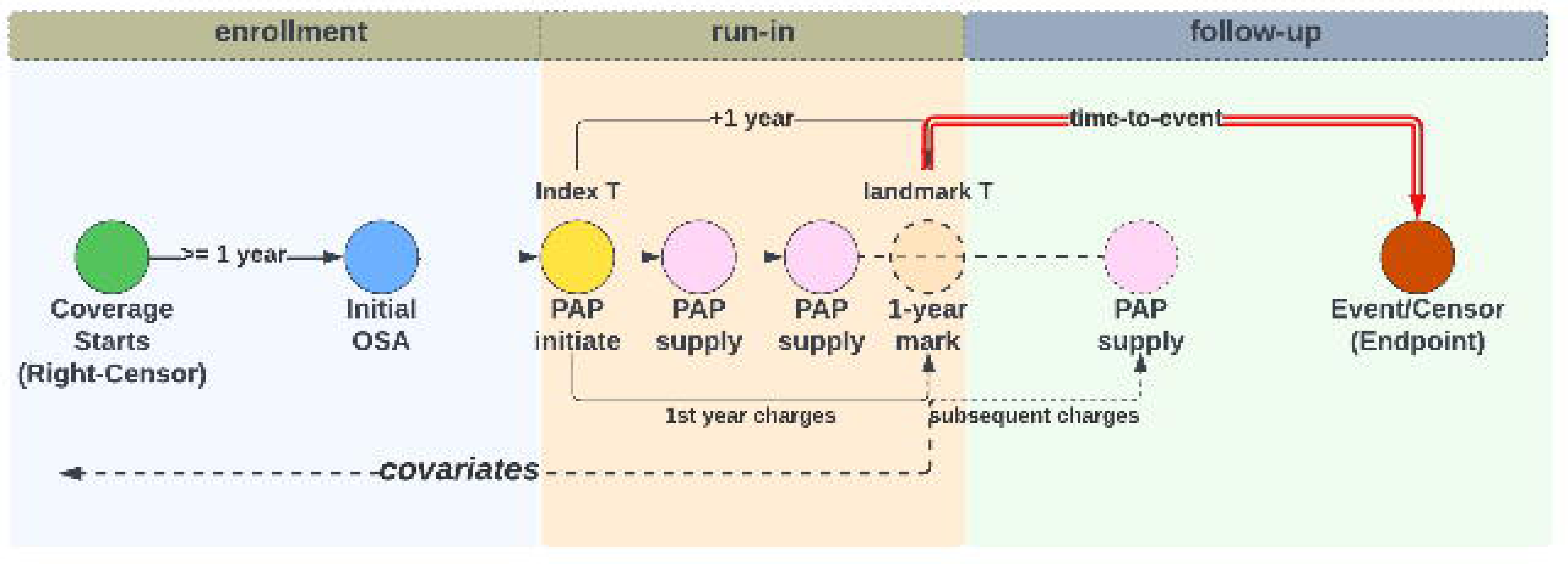
Overview of the PAP utilization exposure group study design.

### PAP utilization definitions

Evidence of PAP initiation was based on the first PAP initiation claim after OSA diagnosis (***Supplemental methods***). PAP utilization exposure groups were used as surrogate measures of PAP adherence. We explored different surrogates based on the total count of PAP claims during first year of utilization. Prior studies have suggested that a greater number of DME claims, such as mask refills, correlates with objective adherence based on hours of use^42^. A comprehensive comparison of all methods is presented in the ***Supplemental methods*** and informed the selection of the quartile-based PAP utilization exposure group definition, which was then used in our primary analysis.

### Study Outcomes

We assessed two primary outcomes: all-cause mortality and incidence of MACE. All-cause mortality was obtained by identifying date of death provided in the Medicare beneficiary summary file. MACE was defined as a composite of first occurrence of myocardial infarction (MI), heart failure (HF), stroke or coronary revascularization, identified by diagnostic and procedure codes (**Supplemental Table 1**). Secondary analyses using each component of MACE as outcomes are also presented.

### Covariates

We included the following covariates: age, sex, race (White, Black, American Indian, Asian, Other), socioeconomic status, prior history of type 2 diabetes, hypertension, obesity, atrial fibrillation, MACE (only in models assessing all-cause mortality), chronic obstructive pulmonary disease (COPD), chronic kidney disease (CKD), anxiety disorder, hypersomnia, insomnia, Charlson comorbidity index (CCI)^43,44^, prescriptions of anticoagulants, antihypertensives, antilipidemic agents and blood glucose regulators (see **Supplemental Table 1** for definitions).

### Statistical analyses

Sociodemographic characteristics and clinical history were described between PAP initiation exposure groups using counts and percentages for categorical data and median and interquartile range for continuous data. Univariate associations between sociodemographic and clinical history variables with PAP initiation exposure groups were performed using chi-squared tests. We used Kaplan-Meier survival analyses and log-rank tests to compare survival curves between PAP initiation exposure groups for MACE and all-cause mortality. We assessed the proportional hazards assumption by assessing scaled Schoenfeld residuals, which did not find strong evidence of violation. We implemented a causal inference framework to determine the effect of PAP initiation or PAP utilization exposure groups on the outcomes of interest (all-cause mortality and MACE. Specific details are presented in the ***Supplemental methods***. Briefly, we derived propensity scores (PS) based on PAP initiation or utilization, and then calculated weights that were used in fully adjusted weighted Cox regression models assessing the effect of PAP initiation or PAP utilization exposure groups on the outcomes of interest, representing causal estimate of the ATE derived from a doubly robust estimator. This framework was applied to our primary analyses, as well as to secondary analyses assessing individual components of MACE. Results of stratified analysis by sociodemographic and clinical characteristics are also presented. We determined statistical significance based on Bonferroni-corrected thresholds of p<0.025 (2 primary outcomes) and provide E-values to assess the strength of association a potential unmeasured confounder would need to express to nullify observed associations^45^.

## Results

### Sample characterization

Our sample included 888,835 eligible Medicare beneficiaries with evidence of OSA diagnosis (median [Q1, Q3] age 73 [69, 78] years; 43.9% women; median [Q1, Q3] follow-up 3 [1.5, 5.1] years). **Figure 3** shows the study flowchart. Among eligible participants, 290,015 (32.6%) had evidence of PAP initiation. **Table 1** shows the sample characteristics according to PAP initiation groups. Participants that have initiated PAP were younger, more likely to be women and White, more likely to have been diagnosed with hypersomnia and insomnia, and less likely to be diagnosed with COPD, type 2 diabetes, hypertension, obesity, atrial fibrillation, CKD, anxiety disorders, and to have had a first MACE at OSA diagnosis. Overall, 5-year cumulative mortality rate was 20.5% (based on total N=888,835) and 5-year cumulative MACE incidence was 41.0% (based on those without MACE at baseline, N=572,072). When stratified by PAP initiation exposure groups, those who initiated PAP had a 5-year cumulative mortality rate and cumulative MACE incidence of 12.4% and 27.4% respectively. Those who did not initiate PAP had a 5-year incidence rates of mortality and MACE event of 17.7% and 30.7%, respectively (p<0.001 for both).

**Figure 3.**
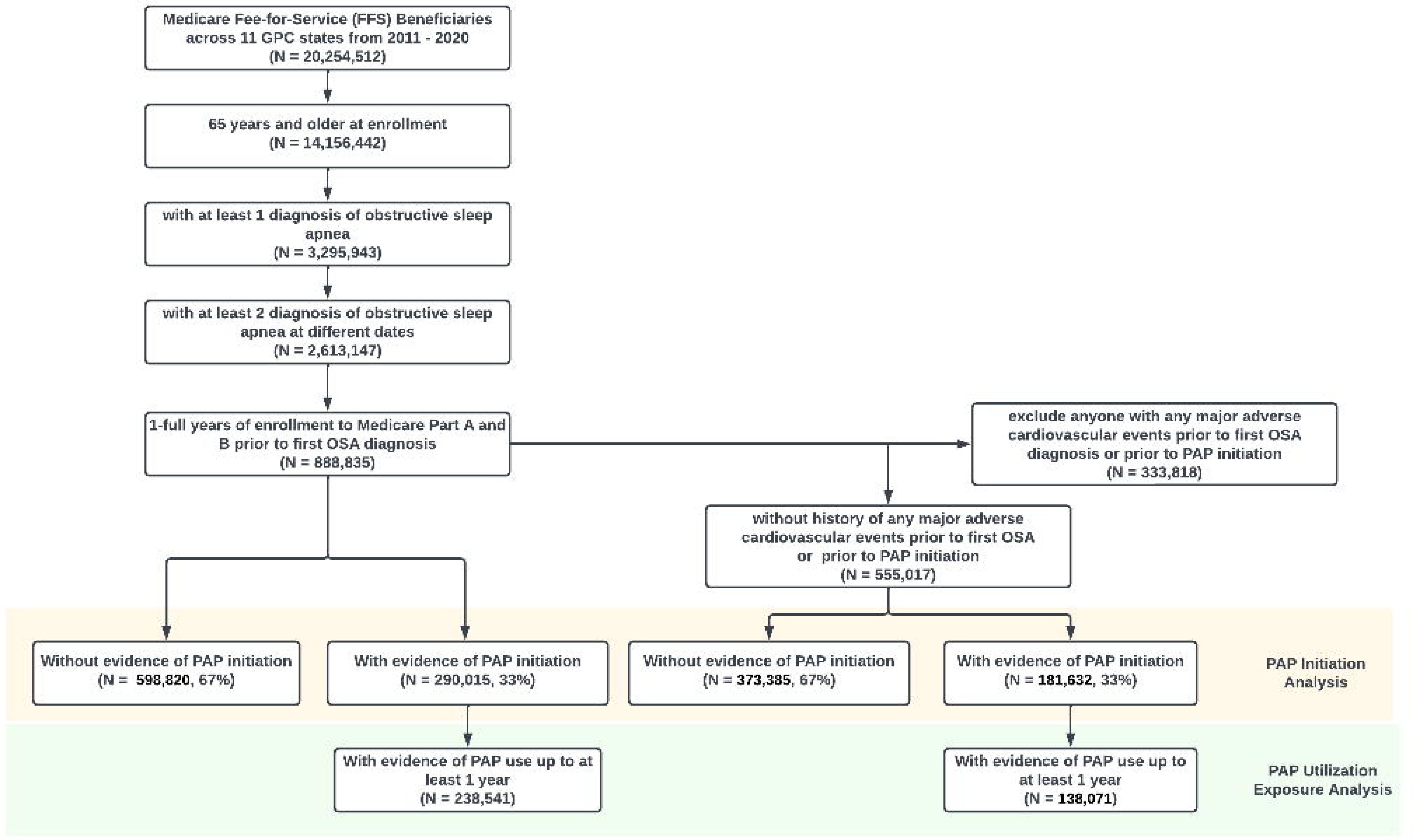
Study cohort definitions and CONSORT diagram.

**Table 1:**
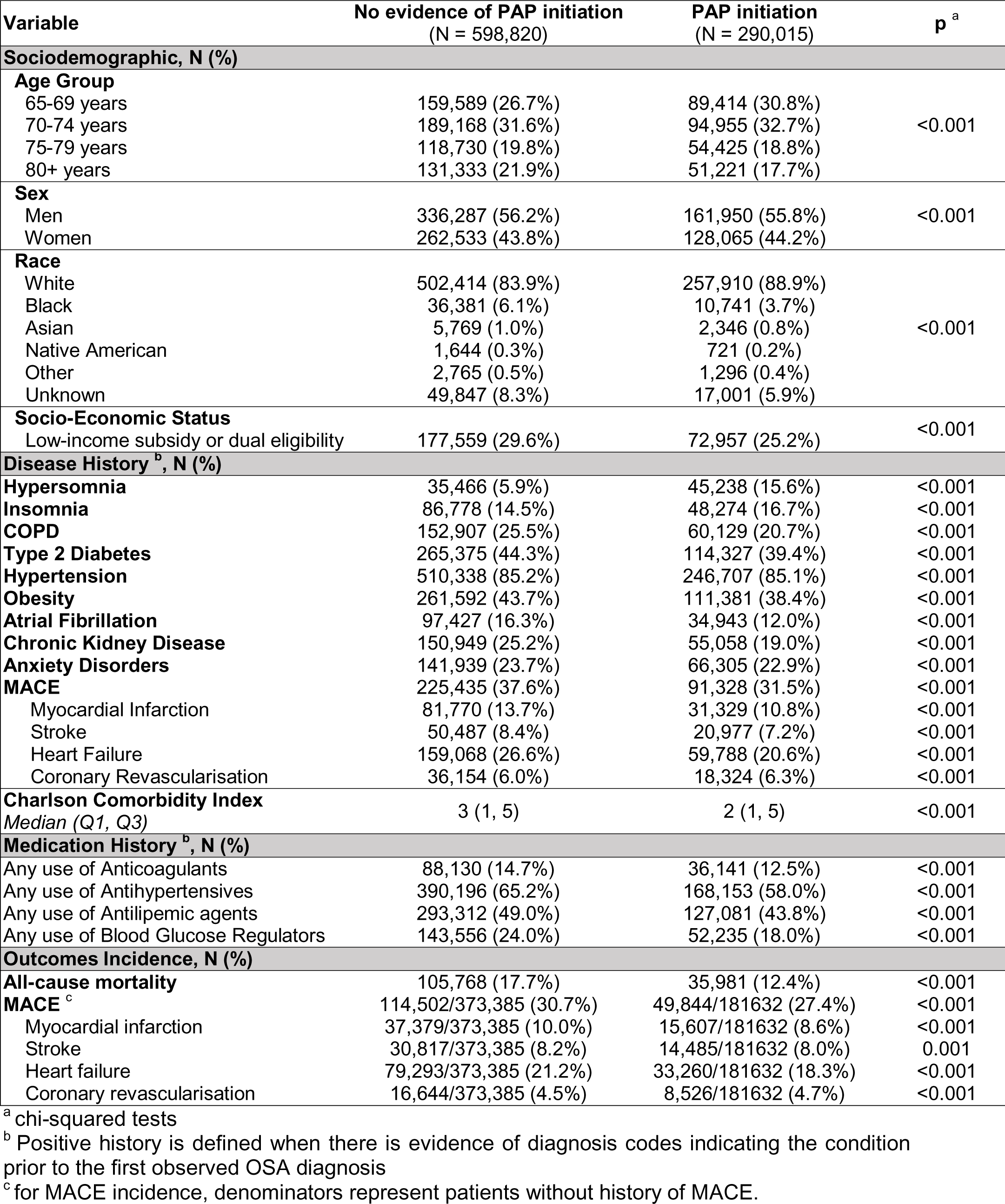
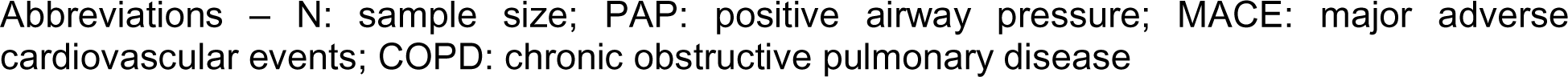
Sample characteristics by positive airway pressure initiation groups among eligible Medicare beneficiaries.

| Variable | No evidence of PAP initiation<br>(N = 598,820) | PAP initiation<br>(N = 290,015) | p<sup>a</sup> |
| --- | --- | --- | --- |
| **Sociodemographic, N (%)** |  |  |  |
| **Age Group** |  |  |  |
| 65-69 years | 159,589 (26.7%) | 89,414 (30.8%) | <0.001 |
| 70-74 years | 189,168 (31.6%) | 94,955 (32.7%) |  |
| 75-79 years | 118,730 (19.8%) | 54,425 (18.8%) |  |
| 80+ years | 131,333 (21.9%) | 51,221 (17.7%) |  |
| **Sex** |  |  |  |
| Men | 336,287 (56.2%) | 161,950 (55.8%) | <0.001 |
| Women | 262,533 (43.8%) | 128,065 (44.2%) |  |
| **Race** |  |  |  |
| White | 502,414 (83.9%) | 257,910 (88.9%) | <0.001 |
| Black | 36,381 (6.1%) | 10,741 (3.7%) |  |
| Asian | 5,769 (1.0%) | 2,346 (0.8%) |  |
| Native American | 1,644 (0.3%) | 721 (0.2%) |  |
| Other | 2,765 (0.5%) | 1,296 (0.4%) |  |
| Unknown | 49,847 (8.3%) | 17,001 (5.9%) |  |
| **Socio-Economic Status** |  |  |  |
| Low-income subsidy or dual eligibility | 177,559 (29.6%) | 72,957 (25.2%) | <0.001 |
| **Disease History<sup>b</sup>, N (%)** |  |  |  |
| Hypersomnia | 35,466 (5.9%) | 45,238 (15.6%) | <0.001 |
| Insomnia | 86,778 (14.5%) | 48,274 (16.7%) | <0.001 |
| COPD | 152,907 (25.5%) | 60,129 (20.7%) | <0.001 |
| Type 2 Diabetes | 265,375 (44.3%) | 114,327 (39.4%) | <0.001 |
| Hypertension | 510,338 (85.2%) | 246,707 (85.1%) | <0.001 |
| Obesity | 261,592 (43.7%) | 111,381 (38.4%) | <0.001 |
| Atrial Fibrillation | 97,427 (16.3%) | 34,943 (12.0%) | <0.001 |
| Chronic Kidney Disease | 150,949 (25.2%) | 55,058 (19.0%) | <0.001 |
| Anxiety Disorders | 141,939 (23.7%) | 66,305 (22.9%) | <0.001 |
| MACE | 225,435 (37.6%) | 91,328 (31.5%) | <0.001 |
| Myocardial Infarction | 81,770 (13.7%) | 31,329 (10.8%) | <0.001 |
| Stroke | 50,487 (8.4%) | 20,977 (7.2%) | <0.001 |
| Heart Failure | 159,068 (26.6%) | 59,788 (20.6%) | <0.001 |
| Coronary Revascularisation | 36,154 (6.0%) | 18,324 (6.3%) | <0.001 |
| **Charlson Comorbidity Index** |  |  |  |
| Median (Q1, Q3) | 3 (1, 5) | 2 (1, 5) | <0.001 |
| **Medication History<sup>b</sup>, N (%)** |  |  |  |
| Any use of Anticoagulants | 88,130 (14.7%) | 36,141 (12.5%) | <0.001 |
| Any use of Antihypertensives | 390,196 (65.2%) | 168,153 (58.0%) | <0.001 |
| Any use of Antilipemic agents | 293,312 (49.0%) | 127,081 (43.8%) | <0.001 |
| Any use of Blood Glucose Regulators | 143,556 (24.0%) | 52,235 (18.0%) | <0.001 |
| **Outcomes Incidence, N (%)** |  |  |  |
| **All-cause mortality** | 105,768 (17.7%) | 35,981 (12.4%) | <0.001 |
| **MACE<sup>c</sup>** | 114,502/373,385 (30.7%) | 49,844/181632 (27.4%) | <0.001 |
| Myocardial infarction | 37,379/373,385 (10.0%) | 15,607/181632 (8.6%) | <0.001 |
| Stroke | 30,817/373,385 (8.2%) | 14,485/181632 (8.0%) | 0.001 |
| Heart failure | 79,293/373,385 (21.2%) | 33,260/181632 (18.3%) | <0.001 |
| Coronary revascularisation | 16,644/373,385 (4.5%) | 8,526/181632 (4.7%) | <0.001 |

<sup>a</sup> chi-squared tests
<sup>b</sup> Positive history is defined when there is evidence of diagnosis codes indicating the condition prior to the first observed OSA diagnosis
<sup>c</sup> for MACE incidence, denominators represent patients without history of MACE.
Abbreviations – N: sample size; PAP: positive airway pressure; MACE: major adverse cardiovascular events; COPD: chronic obstructive pulmonary disease

### PAP initiation is associated with greater survival and MACE-free probability

Kaplan-Meier survival curves for all-cause mortality and MACE, according to PAP initiation exposure groups are presented in **Figure 4**. Log-rank test indicates significant differences in all-cause mortality and MACE-free survival probabilities (p<0.001), with participants that have initiated PAP presenting greater survival than those without.

**Figure 4:**
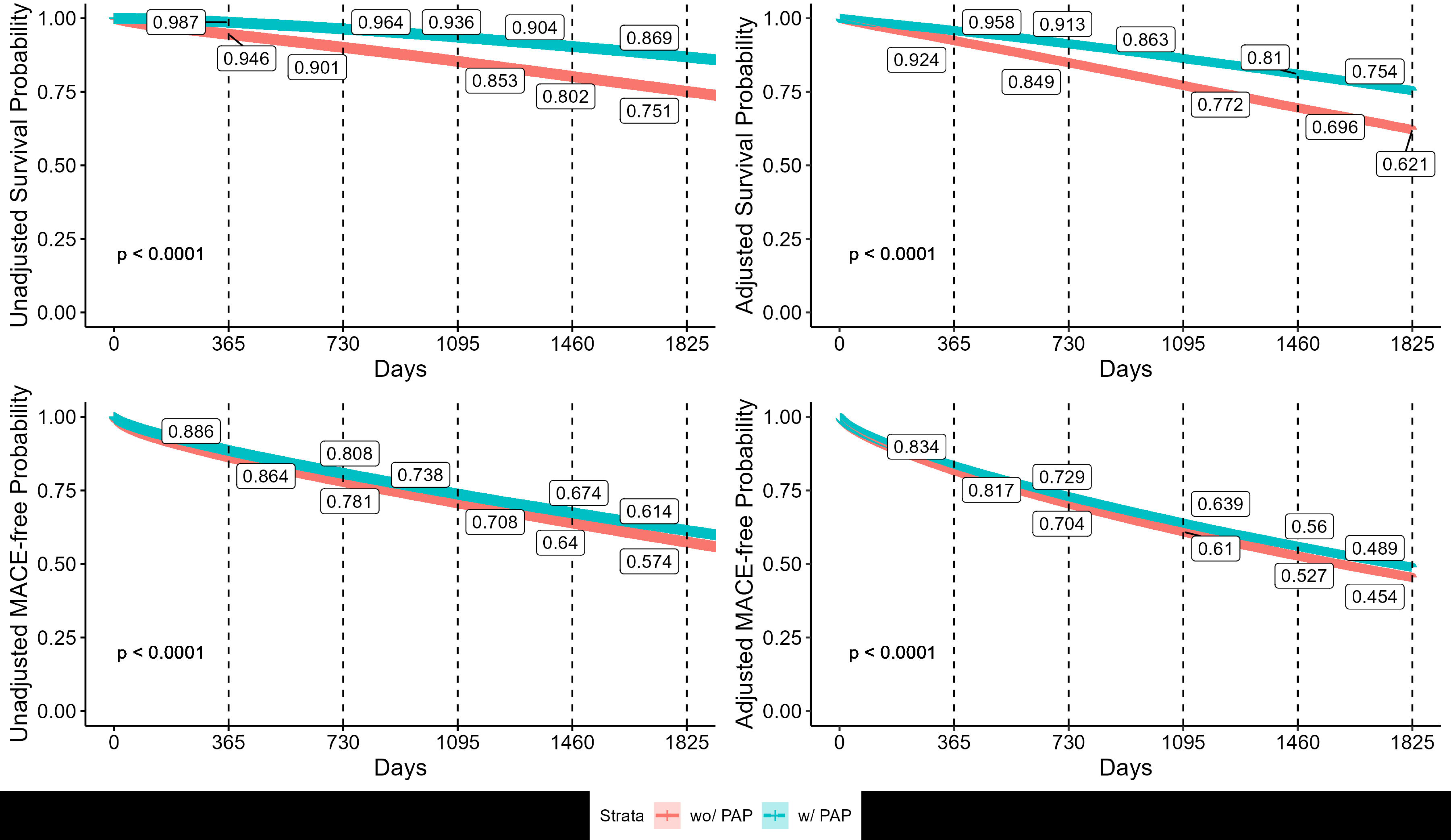
Unadjusted and adjusted Kaplan-Meier survival curves describing the survival probabilities between PAP initiation exposure groups and all-cause mortality (top) and MACE (bottom). Numbers in boxes represent annual survival probabilities per group. The results of log-rank tests comparing exposure groups are also shown.

### PAP initiation is independently associated with lower mortality and lower incidence of MACE

Results of doubly robust Cox proportional hazards models are presented in **Figure 5**. Patients with evidence of PAP initiation had significantly lower all-cause mortality risk (HR [95%CI] = 0.53 [0.52-0.54]; p<0.001) and lower MACE incidence risk (0.90 [0.89-0.91]; p<0.001) when compared to those without evidence of initiating PAP. The risk ratio of an unmeasured confounder (E-value) would need to be 2.47 to explain away the effect of PAP initiation on mortality, and 1.36 to explain away the effect on MACE.

**Figure 5.**
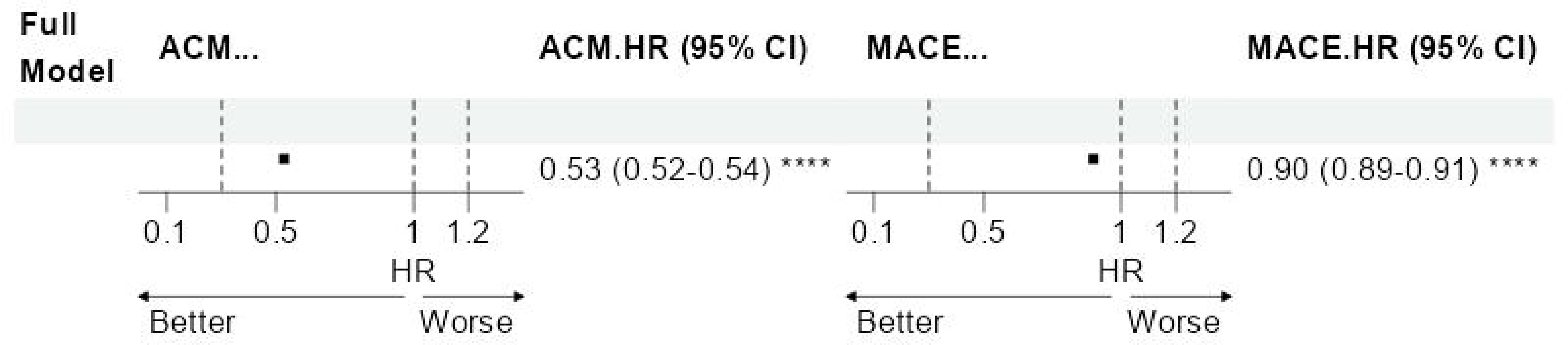
Summary of IPTW-adjusted Cox proportional hazards models assessing the effect of PAP initiation on all-cause mortality (ACM) and MACE in all eligible participants. Results were derived from IPTW-weighted Cox proportional hazards models adjusted for age, sex, race, low-income-subsidy or dual-eligibility indicator, type 2 diabetes, hypertension, obesity, atrial fibrillation, MACE (all-cause mortality only), COPD, CKD, hypersomnia, and insomnia, anxiety disorder, hypersomnia, insomnia, CCI, prescriptions of anticoagulants, antihypertensives, antilipidemic agents and blood glucose regulators. Reference category is no evidence of PAP initiation. Abbreviations: ACM: all-cause mortality; COPD: chronic obstructive pulmonary disease; CKD: chronic kidney disease; CCI: Charlson comorbidity index; PAP: positive airway pressure; HR: hazard ratio; CI: confidence interval; MACE: major adverse cardiovascular event; IPTW: inverse probability of treatment weights. and within categories of relevant sociodemographic and clinical characteristics. Among stratified models, stratification variable was not included as a covariate in propensity score models nor in outcome models.

Analyses of secondary outcomes (MI, HF, stroke, and coronary revascularization) are presented in **Figure 6**. OSA patients with evidence of PAP initiation had significantly lower incidence risk of MI (0.84 [0.82-0.85]; p<0.001; E-value: 1.48), HF (0.89 [0.88-0.90]; p<0.001; E-value: 1.39), and stroke (0.86 [0.84-0.88]; p<0.001; E-value: 1.41). Moreover, PAP initiation was associated with higher incidence risk of revascularization (1.08 [1.05-1.11]; p<0.001).

**Figure 6.**
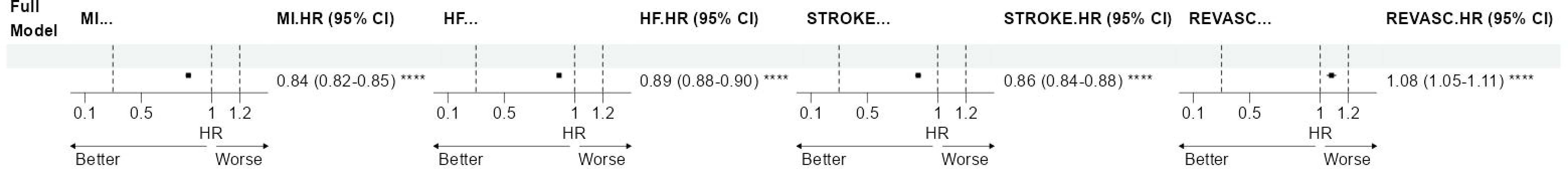
Summary of IPTW-adjusted Cox proportional hazards models assessing the effect of PAP initiation on myocardial infarction (MI), heart failure (HF), stroke (STROKE) and coronary revascularization (REVASC) in all eligible participants. Results were derived from IPTW-weighted Cox proportional hazards models adjusted for age, sex, race, low-income-subsidy or dual-eligibility indicator, type 2 diabetes, hypertension, obesity, atrial fibrillation, COPD, CKD, hypersomnia, and insomnia, anxiety disorder, hypersomnia, insomnia, CCI, prescriptions of anticoagulants, antihypertensives, antilipidemic agents and blood glucose regulators. Reference category is no evidence of PAP initiation. Abbreviations: ACM: all-cause mortality; COPD: chronic obstructive pulmonary disease; CKD: chronic kidney disease; CCI: Charlson comorbidity index; PAP: positive airway pressure; HR: hazard ratio; CI: confidence interval; MACE: major adverse cardiovascular event; IPTW: inverse probability of treatment weights.

Next, we investigated whether the effect of PAP initiation was consistent across subgroups of sociodemographic and clinical factors. Results of primary outcomes are presented in **Supplemental Figure 1** and of secondary outcomes are presented in **Supplemental Figure 2**. We observed that effect estimates regarding all-cause mortality are consistent across subgroups, with significantly lower risk among those that initiated PAP across all strata, and key differences in certain subgroups. We observed stronger protective effects of PAP initiation among women (0.52 [0.51-0.53]) as compared to men (0.53 [0.53-0.54), those with evidence of obesity diagnosis (0.51 [0.50-0.51]) as compared to those without (0.55 [0.54-0.55]), those with evidence of history of atrial fibrillation (0.47 [0.46-0.48]) as compared to those without (0.54 [0.53-0.54]), and those without evidence of hypersomnia (0.52 [0.52-0.53]) as compared to those with (0.63 [0.61-0.65).

Regarding incidence of MACE, we observed stronger PAP initiation effects among those 75 years or older (75-79 years: 0.89 [0.87-0.90]; 80+: 0.86 [0.85-0.88], as compared to those <75 years (65-69 years: 0.91 [0.90-0.93]; 70-74 years: 0.92 [0.90-0.93]). We also observed stronger protective effects of PAP initiation among women (0.88 [0.87-0.89]) as compared to men (0.91 [0.90-0.92]), those with lower social economic status (0.86 [0.85-0.87]) as compared to those with higher (0.91 [0.91-0.92]), those with evidence of insomnia diagnosis (0.87 [0.85-0.89]) as compared to those without (0.90 [0.89-0.91]), those with evidence of obesity diagnosis (0.88 [0.87-0.89]) as compared to those without (0.91 [0.90-0.92]), those with evidence of COPD diagnosis (0.88 [0.86-0.89]) as compared to those without (0.90 [0.90-0.91]), those with evidence of type 2 diabetes diagnosis (0.88 [0.87-0.89]) as compared to those without (0.91 [0.90-0.92]), those with evidence of atrial fibrillation diagnosis (0.85 [0.83-0.87]) as compared to those without (0.90 [0.90-0.91]), and those with higher CCI scores (CCI=1-2 comorbidities: 0.88 [0.87-0.90]; CCI=3-4 comorbidities: 0.89 [0.88-0.91]; CCI=5+ comorbidities: 0.89 [0.87-0.91]) as compared to those with less comorbidities (CCI=0: 0.94 [0.92-0.96]).

Stratified analyses regarding secondary outcomes (MACE components) consistently showed significantly lower incidence of MI, HF, and stroke as well as higher incidence of coronary revascularization procedures across all subgroups of sociodemographic and clinical factors (**Supplemental Figure 2**). Important differences were observed for certain subgroups. Regarding MI, strongest beneficial effects of PAP initiation were observed among women (0.80 [0.79-0.82]), those with lower social economic status (0.80 [0.78-0.82]), those with evidence of hypersomnia diagnosis (0.77 [0.73-0.81]), those with evidence of insomnia diagnosis (0.79 [0.76-0.82]), and those with evidence of anxiety disorder diagnosis (0.79 [0.76-0.82]). Regarding HF, strongest beneficial effects of PAP initiation were observed in those 75 years or older (75-79 years: 0.86 [0.84-0.88]; 80+ years: 0.86 [0.84-0.88]), and those with evidence of atrial fibrillation diagnosis (0.84 [0.81-0.87]). Regarding stroke, strongest beneficial effects of PAP initiation were observed among women (0.83 [0.81-0.85]), those with lower social economic status (0.82 [0.80-0.84]), those with evidence of obesity diagnosis (0.82 [0.80-0.84]), those with evidence of CPOD diagnosis (0.81 [0.79-0.84]), those with evidence of hypertension diagnosis (0.85 [0.83-0.86]), and those with highest CCI scores (0.82 [0.79-0.84]). Regarding coronary revascularization, strongest detrimental effects of PAP initiation were observed among those with 80+ years (1.18 [1.12-1.24]), men (1.10 [1.08-1.13]), and those taking antihypertensive medications (1.12 [1.09-1.15]).

### Determination of first year PAP utilization exposure groups

To inform our proposed analysis assessing the effect of PAP utilization exposure groups, we first observed the distribution of total PAP claims per patient at the end of first year of PAP utilization (**Supplemental Figure 3**). PAP claim counts showed a bimodal distribution, suggesting two utilization patterns were more prevalent: a mode on 3 claims and a mode on 14 claims, which may be suggestive of CMS PAP reimbursement models^46^ and may correlate with PAP adherence^47,48^. Because Medicare claims databases do not include PAP adherence tracking, we explored different PAP exposure group definitions (see *PAP utilization definitions* and **Supplemental Figure 4**) to determine relevant cut-offs in the distribution of PAP claims. We further assessed associations between different PAP exposure group definitions and our primary and secondary outcomes. In general, a greater count of PAP claims during the first year was progressively associated with lower all-cause mortality and lower MACE incidence risk (**Supplemental Figure 5**), reaching a plateau after approximately 15-16 claims. When using the numeric count of PAP claims as our exposure, each additional PAP claim was significantly associated with lower hazards of all-cause mortality (0.98 [0.98-0.98]) and MACE (0.99 [0.99-0.99]). To address the non-linearity of the associations between PAP claim counts and outcomes, we performed spline extrapolation analyses and estimated the HR as a function of PAP claim counts (**Supplemental Figure 6**). Results suggest HR estimates are the highest (e.g., greater hazards) around 4-5 PAP claims and lowest at 17-19 PAP claims across all study outcomes. Taken together, these exploratory analyses helped to inform us that using a *<u>quartile-based definition of PAP exposure groups based on claim counts</u>* (Q1: 1 to 7 claims; Q2: 8 to 12 claims; Q3: 13 to 15 claims; and Q4: >15 claims) provided a realistic representation of PAP utilization patterns that demonstrated clinically relevant variation in incident outcome risk. Therefore, we focused the description of the following results based on this definition.

### Greater PAP utilization is progressively associated with lower risk of mortality and MACE

Results of doubly robust Cox proportional hazards models assessing the effect of PAP utilization exposure groups based on quartiles of PAP claim counts on all-cause mortality and incidence of MACE are presented in **Figure 7** (among all participants) and **Supplemental Figure 7** (stratified by sociodemographic and clinical characteristics), and results for secondary outcomes are presented in **Figure 8 and Supplemental Figure 8**. Among patients with OSA and with evidence of PAP utilization in their first year of therapy, higher quartiles of claim counts (i.e., higher PAP utilization) were progressively associated with lower all-cause mortality (Q2: 0.84 [0.81-0.87], Q3: 0.76 [0.74-0.79], Q4: 0.74 [0.72-0.77]) and lower MACE incidence (Q2: 0.92 [0.89-0.95], Q3: 0.89 [0.86-0.91], Q4: 0.87 [0.85-0.90]), when compared to those with ≤8 PAP claims in their first year. Similar results were observed for secondary outcomes, with lower MI incidence risk on Q3 (0.88 [0.83-0.93]) and Q4 (0.84 [0.79-0.88]), lower HF incidence risk (Q2: 0.91 [0.88-0.95], Q3: 0.86 [0.83-0.89], Q4: 0.85 [0.82-0.88]) and lower stroke incidence risk (Q2: 0.85 [0.80-0.91], Q3: 0.86 [0.81-0.90], Q4: 0.80 [0.76-0.85]). No evidence of association with incident coronary revascularization was observed (**Figure 8**).

**Figure 7.**
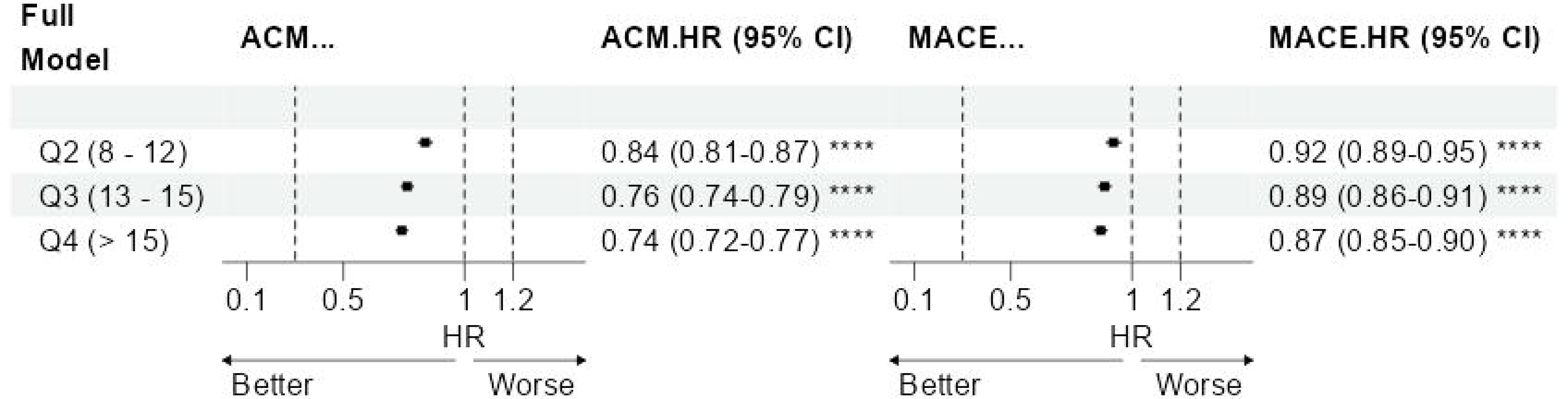
Summary of IPTW-adjusted Cox proportional hazards models assessing the effect of PAP utilization exposure groups based on quartiles of PAP claim counts on all-cause mortality (ACM) and MACE among all participants. PAP exposure groups were defined as follows: Q1 (reference): 1-7 claims; level 2: 8-12 claims; level 3: 13-15 claims; level 4: >15 claims). Results were derived from IPTW-weighted Cox proportional hazards models adjusted for age, sex, race, low-income-subsidy or dual-eligibility indicator, type 2 diabetes, hypertension, obesity, atrial fibrillation, MACE (all-cause mortality only), COPD, CKD, hypersomnia, and insomnia, anxiety disorder, hypersomnia, insomnia, CCI, prescriptions of anticoagulants, antihypertensives, antilipidemic agents and blood glucose regulators. Abbreviations: ACM: all-cause mortality; COPD: chronic obstructive pulmonary disease; CKD: chronic kidney disease; CCI: Charlson comorbidity index; PAP: positive airway pressure; HR: hazard ratio; CI: confidence interval; MACE: major adverse cardiovascular event; IPTW: inverse probability of treatment weights.

**Figure 8.**
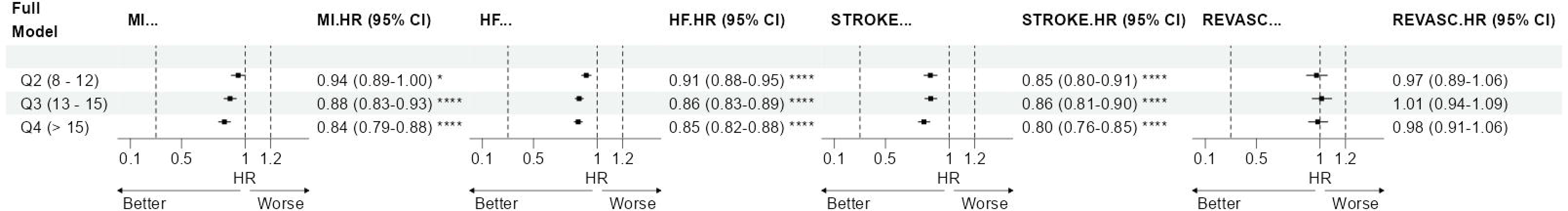
Summary of IPTW-adjusted Cox proportional hazards models assessing the effect of PAP utilization exposure groups based on quartiles of PAP claim counts on myocardial infarction (MI), heart failure (HF), stroke (STROKE) and coronary revascularization (REVASC) in all eligible participants. PAP exposure groups were defined as follows: Q1 (reference): 1-7 claims; level 2: 8-12 claims; level 3: 13-15 claims; level 4: >15 claims). Results were derived from IPTW-weighted Cox proportional hazards models adjusted for age, sex, race, low-income-subsidy or dual-eligibility indicator, type 2 diabetes, hypertension, obesity, atrial fibrillation, COPD, CKD, hypersomnia, and insomnia, anxiety disorder, hypersomnia, insomnia, CCI, prescriptions of anticoagulants, antihypertensives, antilipidemic agents and blood glucose regulators. Abbreviations: ACM: all-cause mortality; COPD: chronic obstructive pulmonary disease; CKD: chronic kidney disease; CCI: Charlson comorbidity index; PAP: positive airway pressure; HR: hazard ratio; CI: confidence interval; IPTW: inverse probability of treatment weights.

Analyses stratified by sociodemographic and clinical characteristics (**Supplemental Figure 7**) were consistent overall, supporting the protective effects of higher PAP claim counts against all-cause mortality and incident MACE across several strata. Exceptions include certain race groups, likely due to the limited sample size of subgroups resulting in wide confidence intervals and less accurate estimates. Regarding all-cause mortality, key differences across strata include stronger protective effects of higher PAP utilization among those with 65-69 years, and those without evidence of hypertension. Regarding MACE, stronger protective effects of higher PAP utilization were observed among those with 65-69 years, those with higher socioeconomic status, those without history of hypertension, those without evidence of atrial fibrillation, and those without evidence of insomnia. Regarding secondary outcomes, (**Supplemental Figure 8**), stronger protective effects of higher PAP utilization were observed against MI among those with 80+ years, those with lower socioeconomic status, and those with evidence of hypersomnia. Regarding HF, stronger protective effects of higher PAP utilization were observed among those with 65-69 years, those without evidence of insomnia, those without evidence of hypertension, and those within less comorbidities according to the CCI. Regarding stroke, protective effects of higher PAP utilization were observed among those without evidence of insomnia. Finally, no consistent associations were observed regarding coronary revascularization procedures. Effects estimates and confidence intervals are shown **Supplemental Figures 7** and **8**.

## Discussion

This study provided a systematic assessment of the effects of PAP initiation and utilization on MACE and mortality among older adults that are Medicare beneficiaries across 11 states in the U.S. Our main findings suggest significant associations between claims-based PAP initiation and utilization and lower all-cause mortality and incidence of MACE. Associations of PAP initiation and utilization were also significant for MACE components, with consistent effects on MI, HF, and stroke. We observed that PAP initiation effects were stronger in women (for both mortality and MACE) in those 75 years or older (MACE only), with lower socioeconomic status (MACE only) and with clinical comorbidities (both mortality and MACE). Higher PAP utilization exposure groups were progressively associated with lower incidence of primary and secondary outcomes, with consistent effects in stratified analysis, but with particularly stronger beneficial effects among those between 65-69 years and those without evidence of hypertension (both mortality and MACE), as well as those with higher socioeconomic status, those without evidence of atrial fibrillation, and those without evidence of insomnia (MACE only). We also found that PAP initiation was associated with higher incidence risk of revascularization, with stronger effects among those 80+ years, men, and those taking antihypertensive medications. We used a robust causal inference framework to minimize immortal time bias and provided more reliable estimates of treatment effects. Results from this observational study might inform future trials assessing the importance of OSA therapy initiation and maintenance towards minimizing CV risk and mortality in older adults.

Extensive epidemiological evidence suggests that moderate-severe OSA is associated with hypertension^49–51^, stroke^26^, atrial fibrillation^52^ MI^53^, and CV mortality^54^. Several mechanisms may explain the relationship between OSA and CV diseases, including sympathetic activation, endothelial dysfunction, oxidative stress, systemic inflammation, cardiac remodeling due to hypoxemia, intrathoracic pressure swings and changes in blood pressure^55^. These overlapping mechanisms indicate that therapies targeting OSA are likely to be beneficial towards CV disease prevention. RCTs have established that PAP improves daytime symptoms, mood, and quality of life^56,57^, and reduces systolic and diastolic blood pressure^49^, especially in patients with resistant hypertension^51^. PAP therapy has positive effects on long-term survival in patients with ischemic stroke and moderate-severe OSA^58^ and it has been associated with lower rate of fatal and non-fatal CV events over a 10-year follow-up^53^.

Despite this consistent evidence, recent RCTs^59–61^ investigating the effect of CPAP on different fatal and non-fatal CV events were negative^27^. These findings were supported by a recent individual meta-analysis of these studies^62^, although a post-hoc on-treatment analysis suggested a protective effect of good adherence to CPAP and reduced risk of MACE^62^. A careful investigation of the limitations in these trials^29^ has suggested that patient selection and treatment adherence may have played an important role on explaining differences between RCTs and observational studies. In fact, less than 20% of patients seeking care in sleep clinics would be eligible to participate in these RCTs^63^. Many patients present with excessive daytime sleepiness, which was an important exclusion criterion in these trials, due to the ethical reason of randomizing patients that are excessively sleepy into control arms^29^. However, we^14,64^ and others^15,65^ have demonstrated this subgroup is at increased risk for incident CV diseases. These observations suggest that alternative approaches using causal inference in well-designed observational studies could provide more generalizable evidence towards the role of PAP therapy on CV outcomes risk.

Exploring ‘real-world data’ approaches using robust statistical methods that minimize biases and confounding issues that are inherent to observational studies may help inform evidence generation in a more timely and cost-effective manner. The success of these approaches has been demonstrated by the current investigation, as it provided relevant and generalizable evidence about the beneficial effects of PAP initiation and utilization in a defined population of older adults in the U.S.. It has also been demonstrated by recent studies exploring the role of CPAP on risk of mortality, HF^35^, stroke^37^, reduced health-care utilisation^66^ and related expenses among those with pre-existing CV diseases^38^, including HF with reduced ejection fraction^67^. In the context of a recently published report from the AHRQ suggesting a lack of strong evidence supporting the role of PAP on long-term CV outcomes^68^, these studies are fundamental to fill this evidence gap.

Our study has identified important associations within subgroups. Notably, we observed greater beneficial effects of PAP initiation on MACE among older adults ≥75 years, suggesting that primary CV prevention among those with newly diagnosed OSA at older ages could be beneficial. However, these results are conflicting with a 3-month RCT of CPAP therapy among elderly adults >70 years with moderate-severe OSA, which did not find significant effects on blood pressure levels or neurocognitive tests, although significant improvements in sleepiness were observed^69^. A secondary analysis of this trial among those >80 years did not find significant effects on sleepiness^70^. Among those that initiated PAP therapy, our study found greater beneficial effects of PAP utilization in those that are 65-69 years. Differences may be explained by length of follow-up and study design considerations. Future trials assessing the role of PAP therapy on health outcomes of older adults using longer follow-up times are warranted. We also observed stronger PAP initiation and greater utilization effects on mortality and MACE among women, in agreement with prior studies, particularly post-menopause^56^. It is likely that women are subjected to stronger acute detrimental effects of OSA following menopause, and therefore might experience stronger therapeutic effects. Women with OSA are also more likely to have comorbid metabolic conditions^71^. However, little is known about sex differences in PAP treatment responses^72^, particularly at older ages. Patients with lower socioeconomic status may be less receptive to receiving PAP therapy^73^ and report poorer treatment adherence^74^. Yet, our results demonstrate that those with low-income subsidy and dual Medicare/Medicaid eligibility have greater benefit of PAP against MACE. These results highlight opportunities to implement care programs designed to minimize health disparities, as recently proposed^75^. Our study also identified stronger effects of PAP initiation among those with comorbid conditions, particularly obesity and atrial fibrillation (for both mortality and MACE), as well as COPD, type 2 diabetes, and those with more comorbidities according to the CCI. It is likely that these subgroups are at a greater underlying risk with the added comorbid impact^12,76–78^, and therefore therapy benefit might be greater. Importantly, subgroups with evidence for comorbidities might also represent patients at highest risk for early termination of CPAP^79^, which suggests that efforts to support therapy continuation might be beneficial particularly for these patients.

We also found important differences among those with prior history of hypersomnia or insomnia. While we found stronger beneficial effects of PAP initiation against mortality among those without evidence of hypersomnia, we observed stronger effects of PAP initiation against MACE among those with evidence of hypersomnia, with results being driven by stronger effects against MI. Results were also corroborated by stronger effects of greater PAP utilization against MI among those with evidence of hypersomnia. These results support the well-established epidemiological relationship between excessive daytime sleepiness and increased CV risk among those with OSA^14,15,64,65^. It is important to note that granular measures of excessive daytime sleepiness might provide a more robust phenotype when compared to claims for hypersomnia. Moreover, only ∼9% of our study cohort had evidence of hypersomnia, which suggests it might not capture all patients with the excessively sleepy phenotype. We also observed stronger effects of PAP initiation against MACE among those with evidence of insomnia. These results also support studies that demonstrate greater CV risk associated with comorbid insomnia and sleep apnea (COMISA). In summary, these results might help guide future RCTs assessing the role of PAP initiation and/or strategies to increase PAP adherence among higher-risk subgroups.

Unexpected results between PAP initiation and increased risk of coronary revascularization procedures, suggesting potentially damaging effects of PAP. Interestingly, these results were mostly driven by older beneficiaries (>75 years). Moreover, analysis of PAP utilization based on claims did not show significant effects of greater PAP utilization and increased risk of revascularization. Future studies assessing the role of PAP initiation on the incident risk of revascularization procedures might provide further insights.

This study utilized state-wide Medicare claims data to determine the effects of PAP initiation and utilization on outcomes. While we acknowledge that objective definitions of PAP adherence and efficacy are preferred to estimate exposure to PAP, telemonitoring data has only become available in recent years^80^. Prior studies have investigated similar PAP exposure definitions in a nationwide claims database in France^35,79^ and within 5% Medicare beneficiaries samples in the U.S.^36–38,81^, and reported consistent results as with objective PAP adherence data^66,67^. From an epidemiologic perspective, state-wide claims databases cover a large proportion of the older adult demographic and offer a scalable approach to identify those likely to be exposed to PAP independently of vendor or telemonitoring capabilities, thus maximizing generalizability. A single-site study comparing objective adherence based on hours of use found that mask refill claims positively correlated with PAP utilization^42^. To provide further information about PAP utilization based on claims, our study presented a detailed assessment of different PAP utilization exposure group definitions. We observed a bimodal distribution of PAP claim counts within the first year since PAP initiation, which is suggestive of recognized CMS reimbursement models ^46^. CMS reimburses PAP therapy among eligible beneficiaries in monthly instalments; when a patient does not meet the eligibility criteria for continued therapy at the third month of utilization, the device is not reimbursed, and no further claims will be identified. We speculate that the first mode of the distribution (3 claims) may represent patients that did not meet the CMS criteria for adherence and therefore may present lower overall PAP utilization during their first year. Similarly, the second mode (14 claims) may represent patients that have demonstrated early adherence and continued to use PAP throughout the year. Our results are consistent with this definition, as it suggests that greater number of PAP claims are progressively associated with greater beneficial outcomes. Therefore, while large scale multisite studies predicting short-and long-term objective PAP adherence using claims data are still needed, the definitions used in our study seem to capture relevant clinical outcomes regarding PAP utilization.

Our study has important limitations. First, our cohort identification approach was based on ICD codes, available through a Medicare claims database, thus preventing characterization of disease severity based on the apnea-hypopnea index (AHI) or other novel physiological traits that have been recently demonstrated to be important for CV risk stratification, such as hypoxic burden^16^ and heart rate responses to events^17^. However, we used an algorithm validated in different clinical sites that demonstrated excellent predictive performance^40^, suggesting that our cohort definition reliably captured individuals with OSA. Second, our claims-based PAP utilization definition may not accurately represent more objective definitions of PAP use. However, hours of use (often used as the only exposure when telemonitoring data is available) may not be the only determinant of PAP efficacy, as other factors such as proportion of protected sleep time (e.g., how many hours sleep on efficacious PAP relative to total sleep time), efficacy of therapy reflected by the optimal pressure, minimal leak, and minimal residual AHI, and even timing of PAP utilization during the night (e.g., early morning termination of therapy might induce more severe rapid eye movement sleep AHI reducing overall PAP efficacy^82,83^), might be more important predictors of successful therapy, and are yet to be consistently assessed in telemonitoring studies. We encourage that PAP vendors expand academic-industry partnerships towards facilitating privacy-preserving linkage of PAP telemonitoring databases with more granular information from EHR, claims, patient reported outcomes and wearables. Such approaches will be fundamental in future studies aimed at determining the effects of PAP therapy on both short-and long-term clinical outcomes. Third, our covariate definition was based only on claims data, and lack granular information about other relevant confounders such as body mass index, blood pressure, and laboratory measurements that are predictive of CV risk. Moreover, other important factors such as diet, physical activity, and healthy adherer behaviours^84^ were not available and could explain some of the observed effects. To mitigate this limitation, we provide E-values as a guide to understand the impact of a potential unmeasured confounders in our reported associations. Results suggest that unmeasured confounders with effects (in the risk ratio scale) as large as 2.47 for all-cause mortality and 1.36 for MACE would have to be observed, after adjustment for other covariates, to explain away associations with our primary outcomes. Finally, the present study focuses on a population of older adults that are Medicare beneficiaries in the Central U.S., and extrapolation of effect estimates to other demographics cannot be made. In one hand, studies in mid-life adults would provide important insights about OSA natural history and early prevention of CV outcomes. On the other hand, our study is generalizable to a very large proportion of older adults, as >95% of those >65 years in the United States are enrolled in Medicare. There is also greater confidence in the ascertainment of outcomes.

In conclusion, PAP utilization based on claims was associated with lower all-cause mortality and MACE incidence (including MI, HF, and stroke) among Medicare beneficiaries >65 years. Results support the hypothesis that PAP has beneficial effects against mortality and CV diseases. This study has the potential to inform future trials assessing the importance of OSA therapy initiation and maintenance towards minimizing adverse health outcomes in older adults leading to longer and healthier lives. These results may also inform strategies to improve adherence and efficacy of PAP therapy there are patient-specific, towards personalized sleep medicine.

## Funding

The study was funded by the American Heart Association (20CDA35310360), Patient-Centered Outcomes Research Institute (RI-CRN-2020-003-IC); NIH CTSA NCATS Frontiers: University of Kansas Clinical and Translational Science Institute (UL1TR002366); Tier 2 grant, University of Missouri.

## Conflict of interest statement

Dr. Krishna Sundar was a past advisory board member for Resmed Inc. and is the co-founder of Hypnoscure LLC through the University of Utah Technology Commercialization Office All other authors declared no conflicts of interest.

## Data availability statement

The data underlying this article cannot be shared publicly to protect the privacy of individuals that participated in the study, as stipulated by each institution of the Greater Plains Collaborative. There are established processes for interested investigators to collaborate with the Greater Plains Collaborative research network, that can be accessed via this URL: https://gpcnetwork.org/.

## Notes

### Greater Plains Collaborative

Sravani Chandaka, Kelechi (KayCee) Anuforo, Lav Patel, Daryl Budine, Nathan Hensel, Siddharth Satyakam, Sharla Smith, Dennis Ridenour, Cheryl Jernigan, Carol Early, Kyle Stephens, Kathy Jurius, Abbey Sidebottom, Cassandra Rodgers, Hong Zhong, Vino Raj, Victor Melendez, Angie Hare, Roman Melamed, Curtis Anderson, Thomas Schouweile, Christine Roering, Philip Payne, Snehil Gupta, John Newland, Albert Lai, Joyce Balls-Berry, Janine Parham, Evin Fritschle, Shanelle Cripps, Kirk Knowlton, Channing Hansen, Erna Serezlic, Benjamin Horne, Jeff VanWormer, Judith Hase, Janet Southworth, Eric Larose, Mary Davis, Laurel Hoeth, Sandy Strey, Brad Taylor, Kris Osinski, April Haverty, Alex Stoddard, Sarah Cornell, Phoenix Do, Lucy Bailey, Beth McDonough, Betsy Chrischilles, Ryan Carnahan, Brian Gryzlak, Gi-Yung Ryu, Katrina Oaklander, Pastor Bruce Hanson, Brad McDowell, Jarrod Field, Abu Mosa, Sasha Lawson, Jim McClay, Soliman Islam, Vasanthi Mandhadi, Kim Kimminau, Dennis Ridenour, Jeff Ordway, Bill Stephens, Russ Waitman, Deandra Cassone, Xiaofan Niu, Lisa Royse, Vyshnavi Paka, Lori Wilcox, Janelle Greening, Carol Geary, Goutham Viswanathan, Jim Svoboda, Jim Campbel, Frances (Annette) Wolfe, Haddy Bah, Todd Bjorklund, Jackson Barlocker, Josh Spuh, Louisa Stark, Mike Strong, Otolose Fahina Tavake-Pas, Rachel Hess, Jacob Kean, Sarah Mumford, Ainsley Huffman, Annie Risenmay, Olivia Ellsmore, Lissa Persson, Kayla Torres Morales, Sandi Stanford, Mahanaz Syed, Rae Schofield, Meredith Zozus, Brian Shukwit, Matthew Decaro, Natalia Heredia, Charles Miller, Alice Robinson, Elmer Bernstam, Fatima Ashraf, Shiby Antony, Juliet Fong Zechner, Philip Reeder, Cindy Kao, Kate Wilkinson, Tracy Greer, Alice Robinson, Lindsay Cowell

## Acknowledgements

The authors would like to acknowledge the technical and scientific resources and support from the Greater Plains Collaborative team members, as well as all Medicare beneficiaries whose data were contributing to this investigation.

## Abbreviations list

ATE: average treatment effect
CCI: Charlson comorbidity index
CI: confidence interval
CMS: Centers for Medicare & Medicaid Services
COPD: chronic obstructive pulmonary disease
CPAP: continuous positive airway pressure
CV: cardiovascular
DME: durable medical equipment
HF: heart failure
HR: hazards ratio
MACE: major adverse cardiovascular events
MI: myocardial infarction
OSA: obstructive sleep apnea
PAP: positive airway pressure
PS: propensity scores
RCT: randomized controlled trials.

## Notes

### Author Declarations

This study was reviewed by the Institutional Review Board of the University of Missouri-Columbia on behalf of each participating institution of the Greater Plains Collaborative (Allina Health, Indiana University, Intermountain Healthcare, Marshfield Clinic, Medical College of Wisconsin, University of Iowa, University of Kansas Medical Center, University of Nebraska Medical Center, University of Texas Health Sciences Center at San Antonio, University of Texas Southwestern, University of Utah, Washington University in St. Louis) and determined to be minimal risk and therefore approved the waiver of authorization under the Privacy Rule. 45 CFR 164.512 (i)(1)(i)

### Summary of Updates

The study incorporated additional Medicare claims data from 2018-2020, substantially increasing the sample size. As a consequence, all reported estimates have been updated, including figures and supplemental materials. The conclusions were very similar with some additional results that are discussed in greater detailed in this version.

