## Supplemental Materials for "Positive Airway Pressure Therapy Predicts Lower Mortality and Major Adverse Cardiovascular Events Incidence in Medicare Beneficiaries with Obstructive Sleep Apnea"

**Supplemental methods**

*Study cohort*

Medicare beneficiaries (>65 years) enrolled to part A and B, and ≥2 distinct obstructive sleep apnea (OSA) claims were collected from multi-state, state-wide, multi-year (2011-2020) Medicare fee-for-service claims data. State-level Medicare claims data were originally obtained as part of the Greater Plains Collaborative Reusable Observable Unified Study Environment (GROUSE)^1^ and the study protocol has been approved by Institutional Review Boards at each participating institution. The Greater Plains Collaborative (GPC) ^2^ is a Patient Centered Outcome Research Network (PCORnet) Clinical Data Research Network including 13 healthcare systems with catchment area across 11 states in the Central U.S.: Kansas, Missouri, Iowa, Wisconsin, Nebraska, Minnesota, Texas, Utah, North Dakota, South Dakota and Indiana.

We defined our study cohort based on a validated EHR algorithm to identify participants with OSA as described by Keenan et al. 2020^3^. Those with two or more claims with International Classification of Diseases (ICD)-9 or 10 codes for OSA at different dates were classified as having OSA (see **Supplemental Table 1**). This algorithm presented optimal predictive performance across six health systems in the U.S., with overall positive predictive value (95%CI) of 97.1% (95.6, 98.2) and negative predictive value of 95.5% (93.5, 97.0)^3^. We further required at least 1-year enrolment with Medicare before their first OSA claim, to better capture beneficiaries that were newly diagnosed with OSA and with complete PAP utilization history since OSA onset. For the analysis of incident MACE, we further excluded beneficiaries with history of any MACE prior to their initial OSA diagnosis date (see *Study Outcomes* for MACE definitions).

*Overview of study design*

This is an observational, retrospective analysis of state-wide Medicare claims data. Due to its observational nature, we proposed two complementary causal inference designs to estimate the average treatment effect (ATE) of PAP initiation (**Figure 1**) and PAP utilization exposure groups based on counts of PAP claims in the first year since initiation (**Figure 2**).

While patients diagnosed with OSA have confirmed their exposure to the deleterious effects of OSA at the date of their diagnosis, there is variability on when they start PAP therapy when prescribed. Therefore, during the time window between OSA diagnosis (*index T*, **Figure 1**) and PAP initiation for those that initiated therapy (*landmark T*, **Figure 1**), these patients share the same exposure than those that never initiated PAP. This misclassification of time intervals between exposure groups may lead to immortal time bias and misestimation of treatment effects^4^. To control for this bias, we used the prescription time-distribution matching method^5^. In this method, the definition of “time zero” (exposure assignment, covariate determination, and start of follow-up) for the PAP initiation group is the corresponding PAP initiation date for each participant (*landmark T*, **Figure 1**). The distribution of number of days from OSA diagnosis to PAP initiation in this group is then used to randomly select and match a time zero for each member of the non-exposed group (*pseudo landmark date*, **Figure 1**). If participants in the non-exposure group have events prior to their pseudo landmark date, they are not included in the analysis, as they would not meet inclusion criteria.

For the analysis of PAP utilization exposure groups, we only included beneficiaries that have been enrolled and have not experienced events at the 1-year mark (*landmark T*, **Figure 2**) since their PAP initiation date. Exposure groups were defined based on the distribution of claim counts during this first year of PAP utilization (see *PAP utilization definitions*). Therefore, the “time zero” for each participant was the date of the first anniversary of PAP initiation.

*PAP utilization definitions*

Evidence of PAP initiation was determined based on the first PAP initiation claim after OSA diagnosis. We used the Current Procedural Terminology 4 and Healthcare Common Procedure Coding System codes to identify PAP claims (see **Supplemental Table 1**). PAP initiation claims were mostly available in the DME claims files and participants were classified as exposed (PAP initiation) and non-exposed (no evidence of PAP initiation throughout the exposure period).

PAP utilization exposure groups are used as surrogate measures of PAP adherence. Conventional definitions of PAP adherence are based on device usage hours and days used and are often used to make reimbursement decisions^6^. Because objective measures of adherence (e.g., hours of PAP use) are not available in Medicare claims databases, we explored different surrogates based on the total count of PAP claims during first year of utilization: a) *Rule-based* definition proposed by Wickwire et al^7^ as: low adherers with < 4 charges, partial adherers with 4-12 charges, and high adherers with >12 charges; b) *Quantile-based*, which segments PAP claim counts based on median, terciles or quartiles; c) *Equally spaced*, which divides PAP claim counts based on incremental number of claims (e.g., 4 or 8 claim increments); d) *Empirical*, by estimating cut-points in the distribution of PAP claim counts using a tree-based discretization algorithm (i.e., a greedy algorithm to iteratively identify the most discriminant cut point for each outcome); and e) *Raw claim counts*, representing the numerical PAP claim counts in the first year. Prior studies have suggested that a greater number of DME claims, such as mask refills, correlates with objective adherence based on hours of use^8^. A comprehensive comparison of all methods is presented in the online supplement and informed the selection of the quartile-based PAP utilization exposure group definition, which was then used in our primary analysis.

*Study Outcomes*

We assessed two primary outcomes: all-cause mortality and incidence of MACE. All-cause mortality was obtained by identifying date of death provided in the Medicare beneficiary summary file. The main sources of death information CMS uses are: Social Security Death Master File, Medicare Common Working File and Online date of death submitted by family members^9^. MACE was defined as a composite outcome defined as the first occurrence of myocardial infarction (MI), heart failure (HF), stroke or coronary revascularization, identified by diagnostic and procedure code claims, as specified in **Supplemental Table 1***.* In analyses using MACE as the outcome, we excluded participants with evidence of MACE prior to the index date. Secondary analyses using each individual component of MACE as outcomes were also presented.

*Covariates*

We included the following covariates of interest: age at the first OSA diagnosis, sex, race (White, Black, American Indian, Asian, Other), low-income subsidy or dual-eligibility indicator (determined as the patient being eligible for dual enrolment to Medicaid or Low Income Subsidy for at least 1 month during their enrolment period) representing proxies of socioeconomic status, prior history of type 2 diabetes, hypertension, obesity, atrial fibrillation, MACE (only in models assessing all-cause mortality), chronic obstructive pulmonary disease (COPD), chronic kidney disease (CKD), anxiety disorder, hypersomnia, insomnia, Charlson comorbidity index (CCI, categorized as 0, 1-2, 3-4 or 5+ comorbidities)^10,11^, prescriptions of anticoagulants, antihypertensives, antilipidemic agents and blood glucose regulators. **Supplemental Table 1** describes specific diagnosis and procedure codes utilized to define each computable phenotype above used as covariates.

*Statistical analyses*

Sociodemographic characteristics and clinical history were described between PAP initiation exposure groups using counts and percentages for categorical data and median and interquartile range for continuous data. Univariate associations between demographic and clinical history variables with PAP initiation exposure groups were performed using chi-squared tests. We used Kaplan-Meier survival analyses and log-rank tests to compare survival curves between PAP initiation exposure groups for MACE and all-cause mortality. We assessed the proportional hazards assumption by assessing scaled Schoenfeld residuals, which did not find strong evidence of violation of the proportionality assumption. We implemented a causal inference framework to determine the effect of PAP initiation or PAP utilization exposure groups on the outcomes of interest (all-cause mortality and MACE). A detailed theoretical framework of causal inference in the context of PAP exposures groups can be found elsewhere^12^. First, we derived propensity scores (PS) from regularized generalized regression models using either PAP initiation (logistic) or PAP utilization exposure groups (Poisson or ordinal, depending on definition) as the outcome with a penalizing term tuned using 5-fold cross-validation. Next, we calculated weights based on PS for individuals in each exposure group as 1/PS for the exposed group and 1/(1-PS) for the non-exposed group. These weights were used in fully adjusted weighted Cox regression models assessing the effect of PAP initiation or PAP utilization exposure groups on the outcomes of interest, representing causal estimate of the ATE derived from a doubly robust estimator. This framework was applied to our primary analyses, as well as to secondary analyses assessing individual components of MACE. Competing risk analyses using the Fine-Gray models were also assessed and results were not different from Cox proportional hazards model. Results of analysis stratified by age groups (65-69, 70-74, 75-79, 80+ years), sex, race, low-income subsidy or dual-eligibility, type 2 diabetes, hypertension, obesity, atrial fibrillation, MACE (for all-cause mortality), COPD, CKD, anxiety disorder, hypersomnia, insomnia, CCI categories, prescriptions of anticoagulants, antihypertensives, antilipidemic agents, and blood glucose regulators are also presented. Spline extrapolation analyses were used to represent hazard ratios as a function of total PAP claim counts during first year. We determined statistical significance based on Bonferroni-corrected thresholds of p<0.025 (2 primary outcomes). To determine the strength of association a potential unmeasured confounder would need to express with both the exposure and the outcome that could explain away the observed associations, we calculated E-values relative to effect estimates of our primary and secondary outcomes, as reported by WanderWeele and Ding^13^, using the web application available from Mathur et al.^14^. All data extraction, processing and analyses were conducted using Snowflake SQL and R (version 4.3.0).

**Supplemental Table 1:** Definitions of computable phenotypes used for study exposure, outcomes, and covariates.

| **Computable phenotype** | **Definition** |
| --- | --- |
| Obstructive sleep apnea | ICD-9-CM: 327.20, 327.23, 327.29, 780.51, 780.53, 780.57 |
|  | ICD-10-CM: G47.30, G47.33, G47.39 |
|  | Algorithm: two or more instances at different dates, first instance used to define onset |
| PAP initiation | CPT-4: 94660  HCPCS: E0601, E0470, E0471 |
| PAP utilization | CPT-4: 94660  HCPCS: E0601, E0470, E0471, A4604, A7027, A7028, A7029, A7030, A7031, A7032, A7033, A7034, A7035, A7036, A7037, A7038, A7039, A7044, A7046, E0561, E0562  Algorithm: count distribution used to determine PAP utilization groups |
| Myocardial infarction | ICD-9-CM: 410.X, 412.X |
|  | ICD-10-CM: I21.X, I22.X, I23.X |
|  | Algorithm: first instance after time zero |
| Stroke | ICD-9-CM: 431.X, 434.X |
|  | ICD-10-CM: I61.X, I62.X, I64.X |
|  | Algorithm: first instance after time zero |
| Heart failure | ICD-9-CM: 428.X |
|  | ICD-10-CM: I50.X |
|  | Algorithm: first instance after time zero |
| Cardiac revascularization | ICD-9-CM: 327.20, 327.23, 327.29, 780.51, 780.53, 780.57 |
|  | ICD-10-PCS: 021X, 027X |
|  | HCPCS: 92920-92944, 92973, 92974, 92975, 92980, 92981, 92982, 92984, 92995, 92996 |
|  | Algorithm: first instance after time zero |
| Peripheral procedures | ICD-9-CM: 00.4, 00.66 |
|  | HCPCS: 37220-37235, 37215-37218, 37236-37249, 37211-37214, 37184-37188 |
|  | Algorithm: first instance after time zero |
| Obesity | ICD-9-CM: 278.00, 278.01, 278.03 |
|  | ICD-10-CM: E66.X, Z68.3, Z68.4 |
|  | Algorithm: coded at or prior time zero |
| Type 2 diabetes | ICD-9-CM: 250.X, 357.2, 362.0 |
|  | ICD-10-CM: E10.X, E11.X, E08.42, Z13.42 |
|  | Algorithm: coded at or prior index date |
| Hypertension | ICD-9-CM: 401.X, 402.X, 403.X, 404.X, 405.X |
|  | ICD-10-CM: I10.X, I11.X, I12.X, I13.X, R03.X |
|  | Algorithm: coded at or prior time zero |
| Atrial fibrillation | ICD-9-CM: 427.3  ICD-10-CM: I48.X  Algorithm: coded at or prior time zero |
| Anxiety disorder | ICD-9-CM: 300.X  ICD-10-CM: N18.X  Algorithm: coded at or prior time zero |
| Chronic kidney disease | ICD-9-CM: 585.X  ICD-10-CM: F40.X-F48.X  Algorithm: coded at or prior time zero |
| Hypersomnia | ICD-9-CM: 327.1 |
|  | ICD-10-CM: F51.1, G47.1 |
|  | Algorithm: coded at or prior time zero |
| Insomnia | ICD-9-CM: 327.0, 307.41, 307.42, 780.52 |
|  | ICD-10-CM: F51.01, F51.02, F51.09 |
|  | Algorithm: coded at or prior time zero |
| Chronic obstructive pulmonary disease | ICD-9-CM: 496.X  ICD-10-CM: J44.X  Algorithm: coded at or prior time zero |
| Medications | See: <https://raw.githubusercontent.com/RWD2E/phecdm/main/res/valueset_autogen/ecqm-medication.json> |

Abbreviations: ICD: International Classification of Diseases; CM: Clinical Modification; CPT: Current Procedural Terminology; PCS: Procedure Coding System; HCPCS: Healthcare Common Procedure Coding System

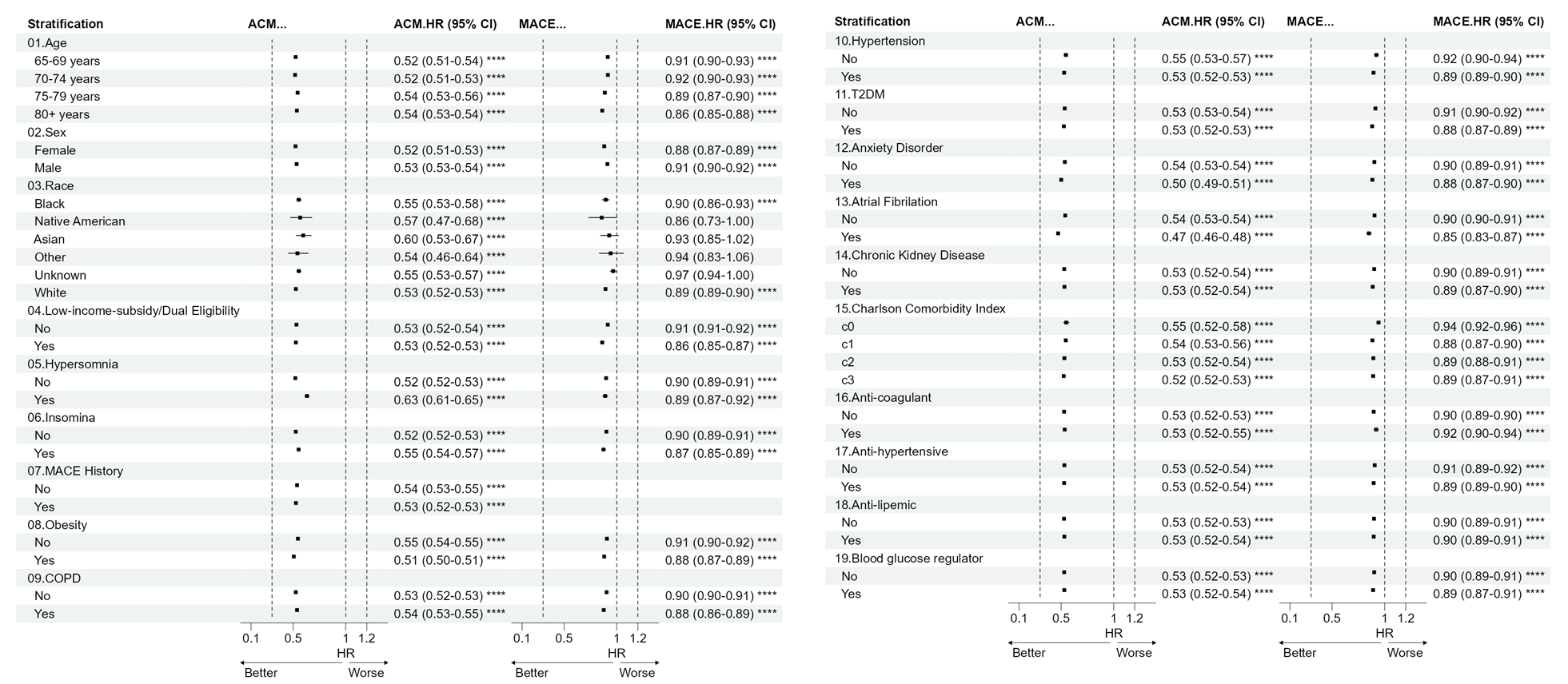

**Supplemental Figure 1.** Summary of inverse probability of treatment weights (IPTW)-adjusted Cox proportional hazards models assessing the effect of PAP initiation on all-cause mortality (ACM) and MACE within categories of relevant sociodemographic and clinical characteristics. Results were derived from IPTW-weighted Cox proportional hazards models adjusted for age, sex, race, low-income-subsidy or dual-eligibility indicator, type 2 diabetes, hypertension, obesity, atrial fibrillation, MACE (all-cause mortality only), COPD, CKD, hypersomnia, and insomnia, anxiety disorder, hypersomnia, insomnia, CCI, prescriptions of anticoagulants, antihypertensives, antilipidemic agents and blood glucose regulators. Among stratified models, stratification variable was not included as a covariate in propensity score models nor in outcome models. Reference category is no evidence of PAP initiation. Abbreviations: ACM: all-cause mortality; COPD: chronic obstructive pulmonary disease; CKD: chronic kidney disease; CCI: Charlson comorbidity index; PAP: positive airway pressure; HR: hazard ratio; CI: confidence interval; MACE: major adverse cardiovascular event; IPTW: inverse probability of treatment weights.

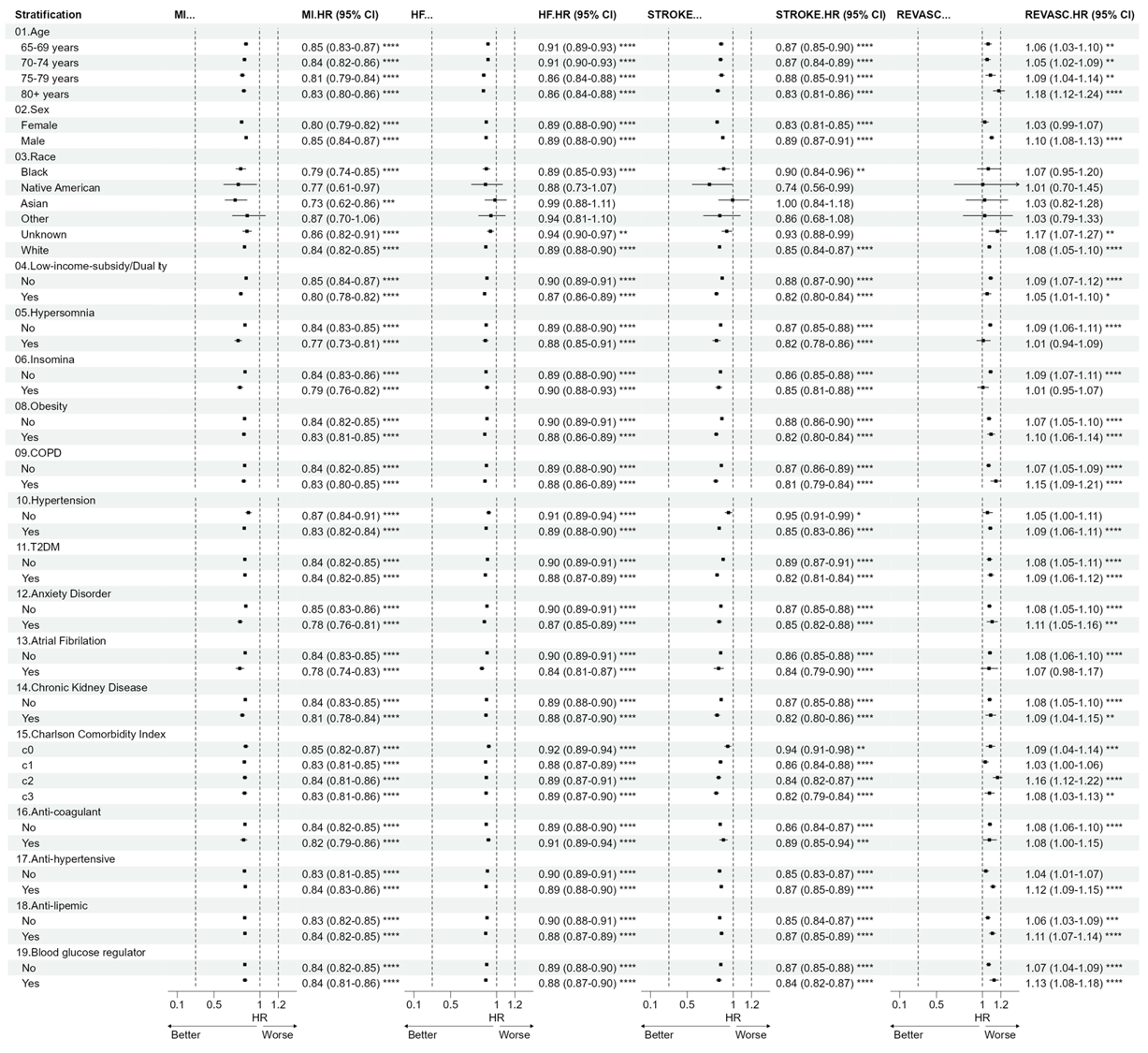

**Supplemental Figure 2:** Summary of inverse probability of treatment weights (IPTW)-adjusted Cox proportional hazards models assessing the effect of PAP initiation on myocardial infarction (MI), heart failure (HF), stroke (STROKE) and coronary revascularization (REVASC) within categories of relevant sociodemographic and clinical characteristics. Results were derived from IPTW-weighted Cox proportional hazards models adjusted for age, sex, race, low-income-subsidy or dual-eligibility indicator, type 2 diabetes, hypertension, obesity, atrial fibrillation, MACE (all-cause mortality only), COPD, CKD, hypersomnia, and insomnia, anxiety disorder, hypersomnia, insomnia, CCI, prescriptions of anticoagulants, antihypertensives, antilipidemic agents and blood glucose regulators. Among stratified models, stratification variable was not included as a covariate in propensity score models nor in outcome models. Reference category is no evidence of PAP initiation. Abbreviations: ACM: all-cause mortality; COPD: chronic obstructive pulmonary disease; CKD: chronic kidney disease; CCI: Charlson comorbidity index; PAP: positive airway pressure; HR: hazard ratio; CI: confidence interval; MACE: major adverse cardiovascular event; IPTW: inverse probability of treatment weights.

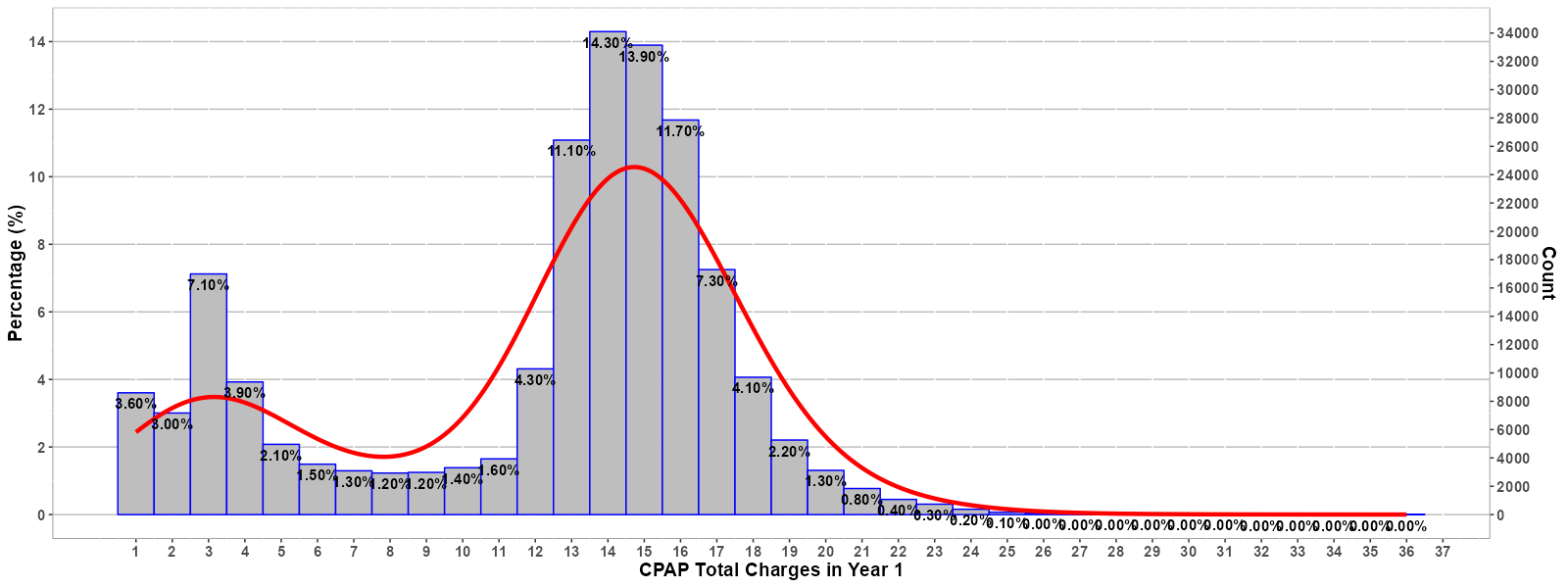

**Supplemental Figure 3.** Distribution of total count of positive airway pressure (PAP) claims during the first year of PAP utilization.

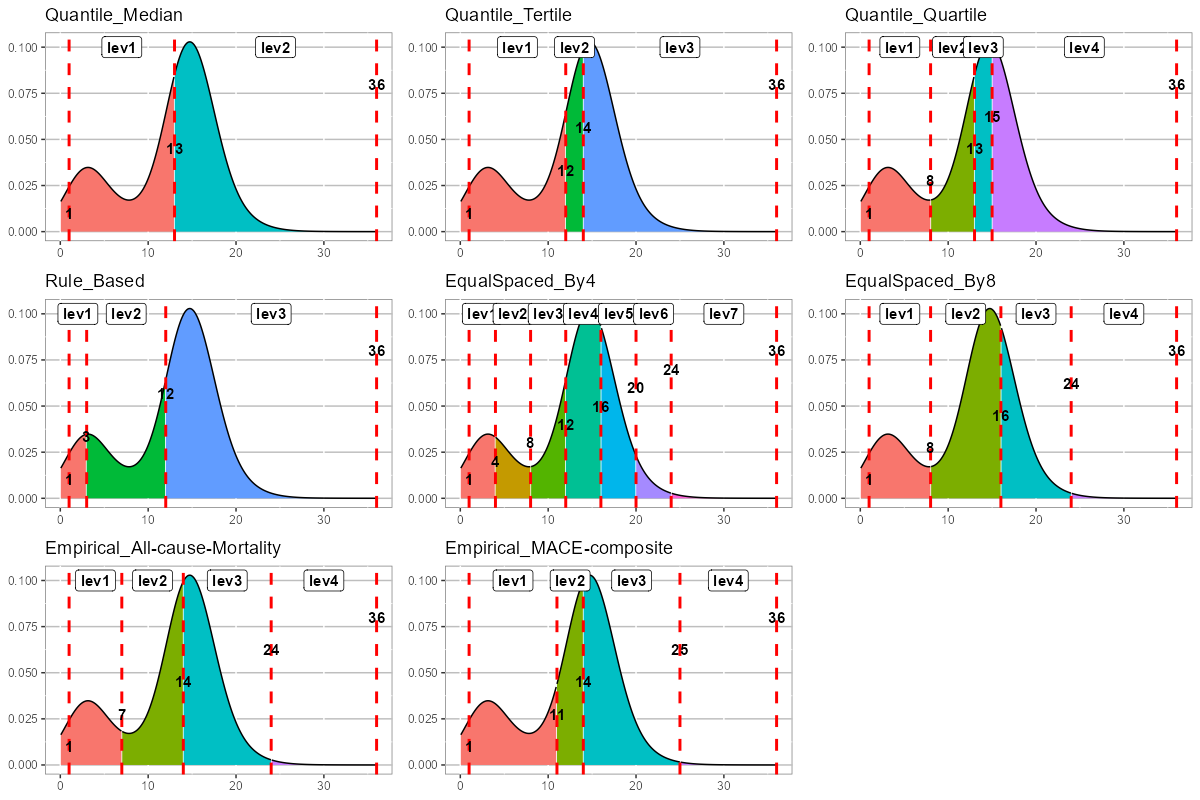

**Supplemental Figure 4.** Categorization of positive airway pressure (PAP) claim counts distribution during first year of PAP utilization according to quantile-based definitions (Quantile_Median, Quantile_Tertile, Quantile_Quartile), rule-based definitions (Rule_Based), equal spaced (EqualSpaced_By4, EqualSpaced_By8) and based on an empirical method that identified the most discriminant cut point for all-cause mortality and MACE (Empirical_All-case-Mortality, Empirical_MACE-composite). Vertical red dashed lines represent cut points for each definition.

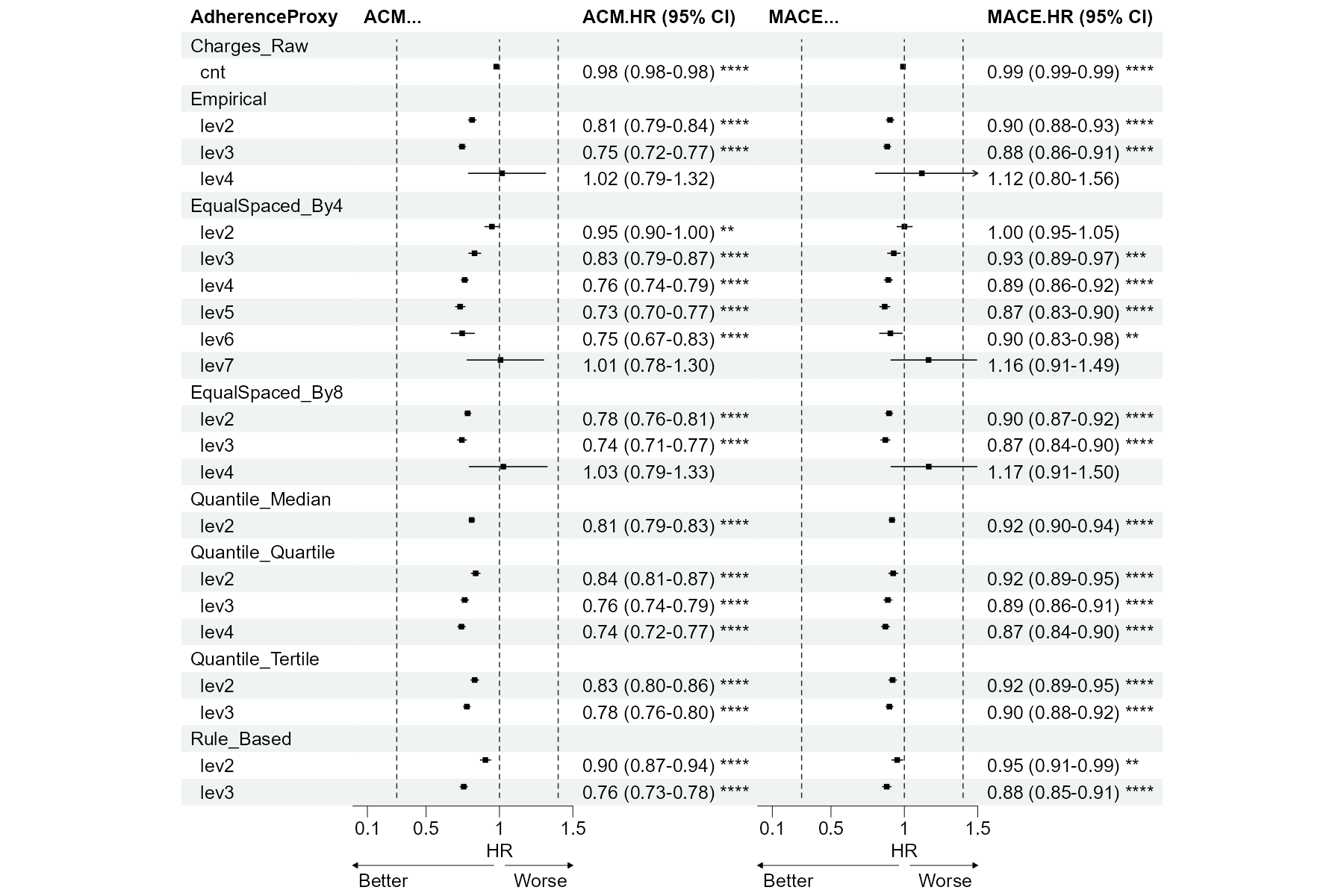
**Supplemental Figure 5:** Summary of inverse probability of treatment weights (IPTW)-adjusted Cox proportional hazards models assessing the effect of different PAP exposure group definitions at first year of PAP utilization on primary outcomes: all-cause mortality (ACM) and major adverse cardiovascular events (MACE).

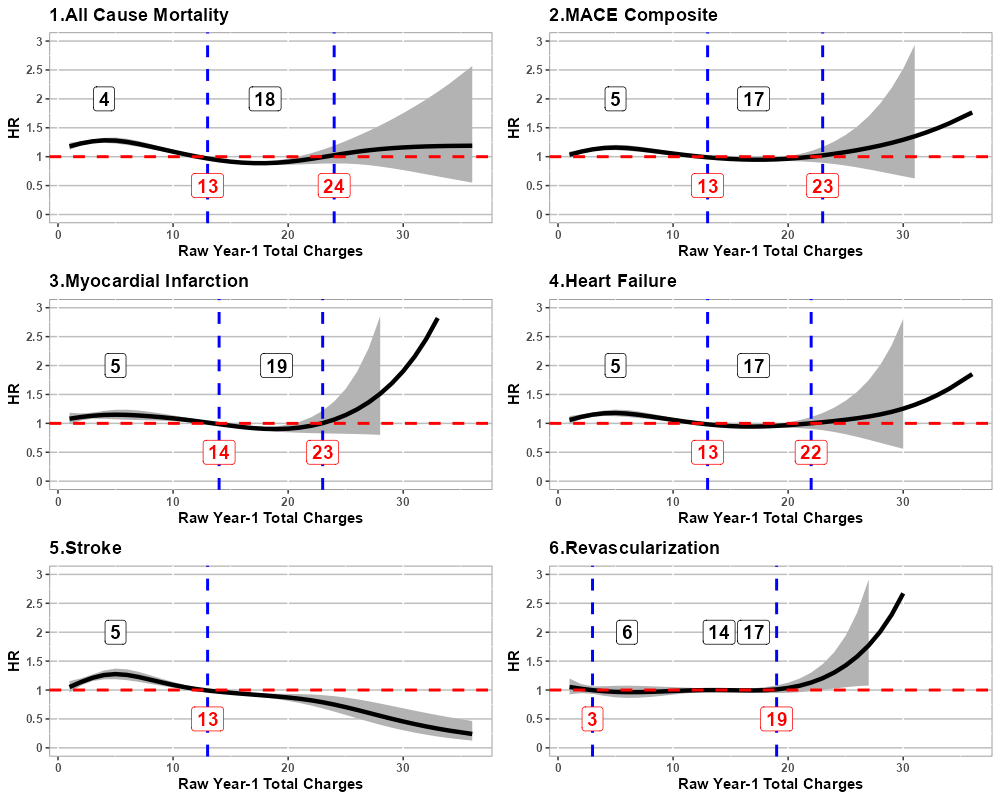

**Supplemental Figure 6:** Spline extrapolation analyses representing estimated hazards ratio (HR) as a function of the total positive airway pressure (PAP) claim counts during first year of PAP utilization for each primary and secondary study outcome. Blue vertical dashed lines represent HR inflection points, with the corresponding count of PAP claims. Number in black boxes represent the number of claims with highest and lowest HR within each inflection. Numbers in red boxes represent the number of claims at the inflection. Red horizontal dashed lines represent the null (HR=1).

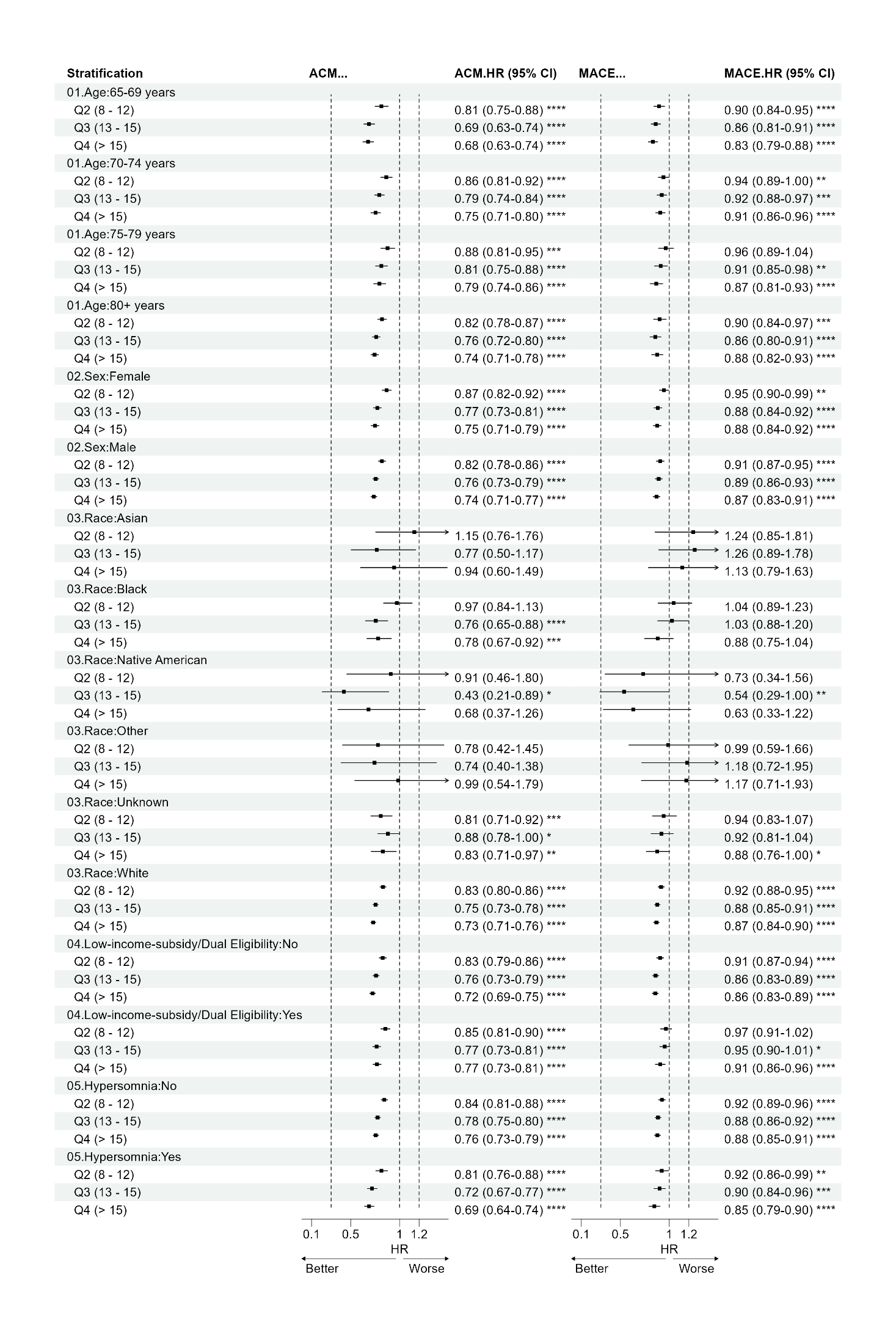

**(continued)**
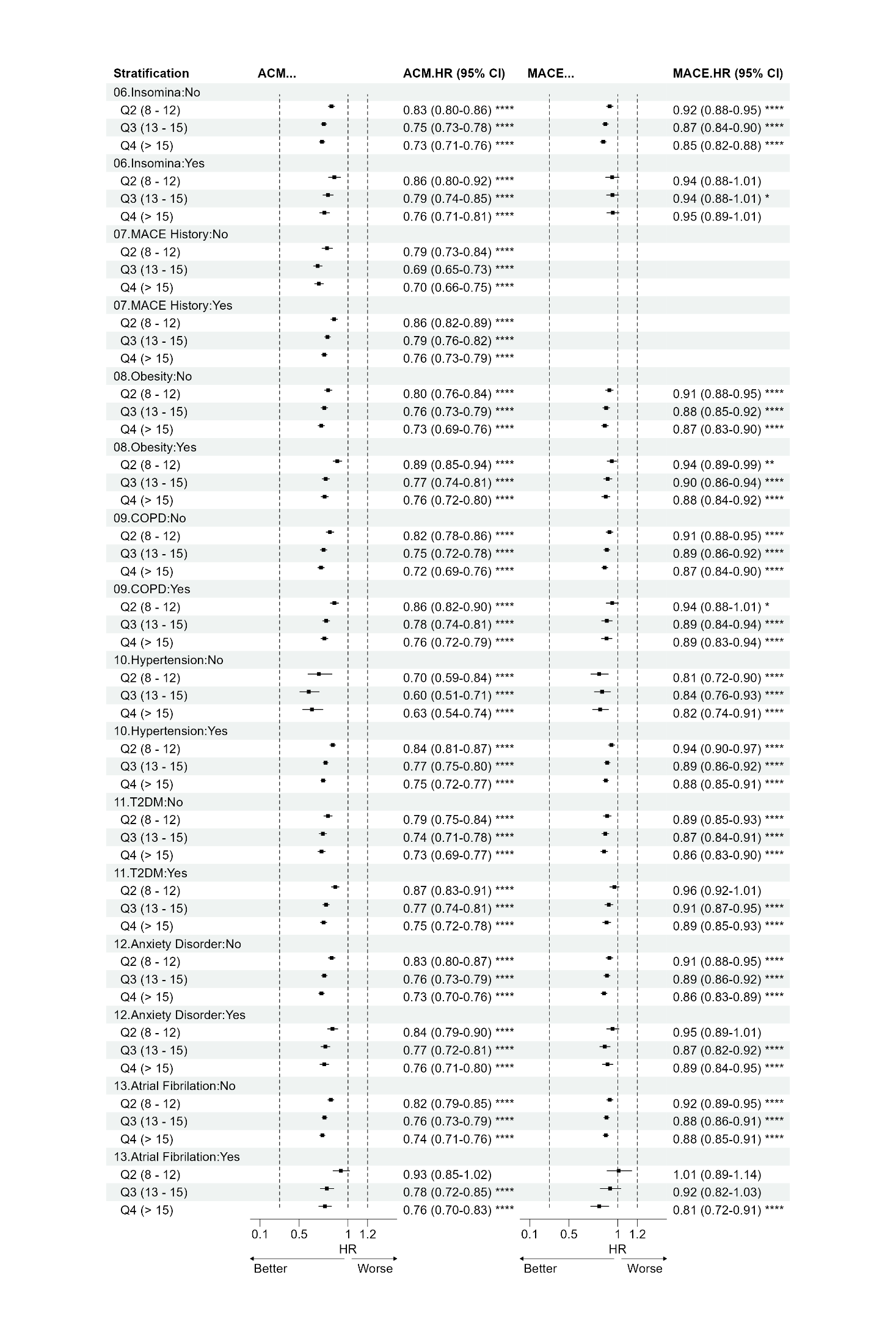

**(continued)**

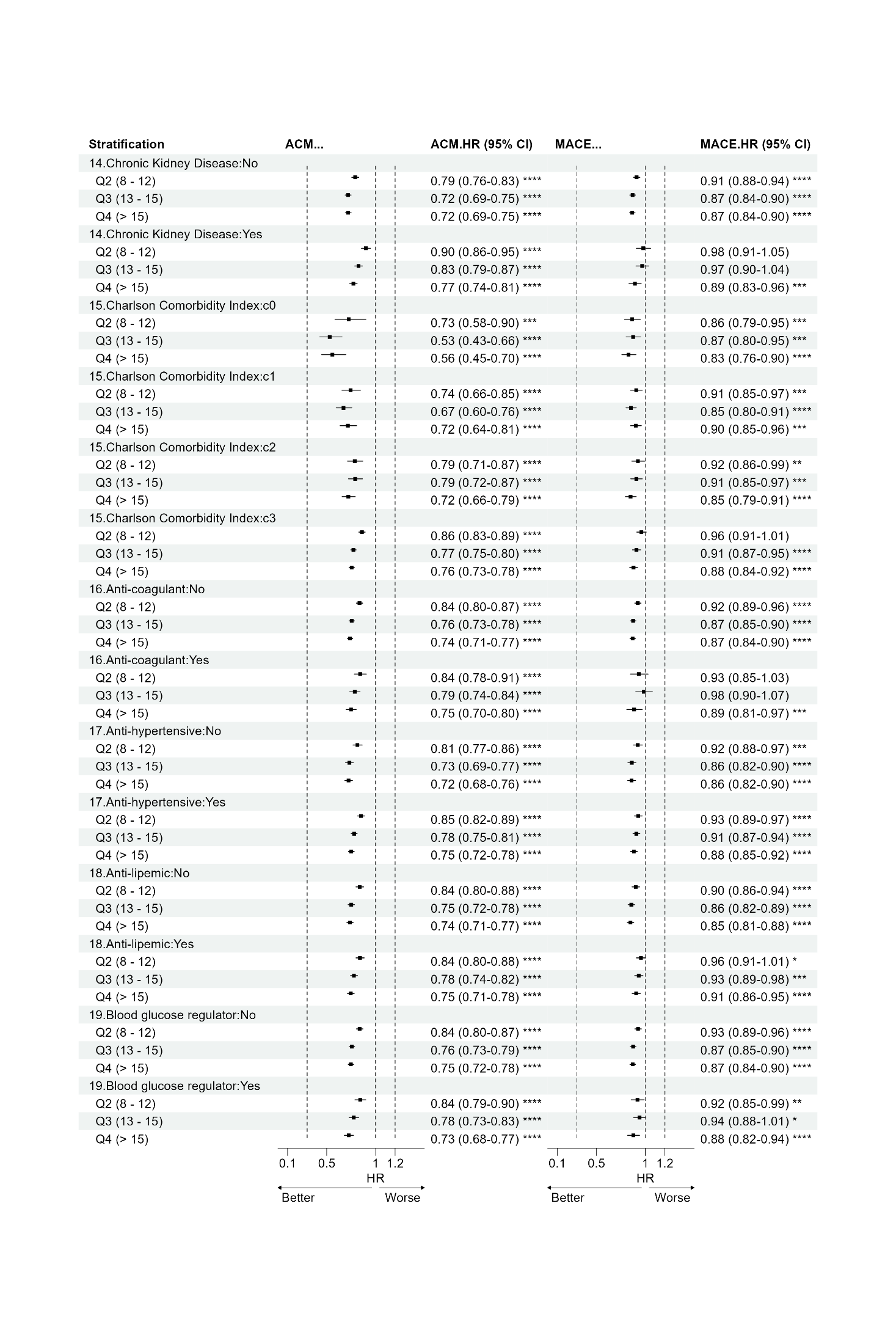

**Supplemental Figure 7:** Summary of inverse probability of treatment weights (IPTW)-adjusted Cox proportional hazards models assessing the effect of PAP exposure groups based on quartiles of PAP utilization during first year on all-cause mortality (ACM) and MACE within categories of relevant sociodemographic and clinical characteristics. PAP exposure groups were defined as follows: level 1 (reference): 1-7 claims; level 2: 8-12 claims; level 3: 13-15 claims; level 4: >15 claims). Results were derived from IPTW-weighted Cox proportional hazards models adjusted for age, sex, race, low-income-subsidy or dual-eligibility indicator, type 2 diabetes, hypertension, obesity, atrial fibrillation, MACE (all-cause mortality only), COPD, CKD, hypersomnia, and insomnia, anxiety disorder, hypersomnia, insomnia, CCI, prescriptions of anticoagulants, antihypertensives, antilipidemic agents and blood glucose regulators. Among stratified models, stratification variable was not included as a covariate in propensity score models nor in outcome models. Reference category is no evidence of PAP initiation. Abbreviations: ACM: all-cause mortality; COPD: chronic obstructive pulmonary disease; CKD: chronic kidney disease; CCI: Charlson comorbidity index; PAP: positive airway pressure; HR: hazard ratio; CI: confidence interval; MACE: major adverse cardiovascular event.

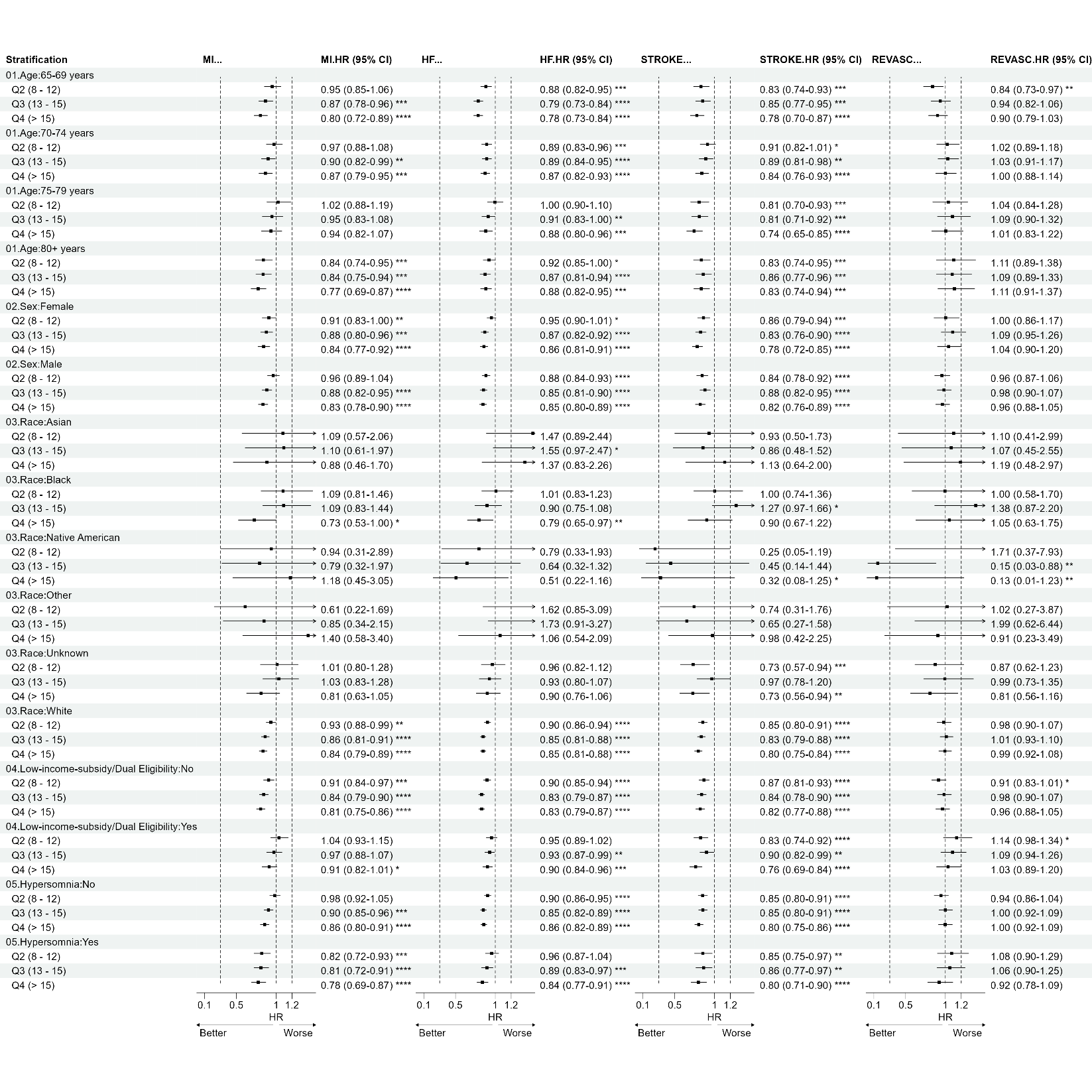

**(continued)**

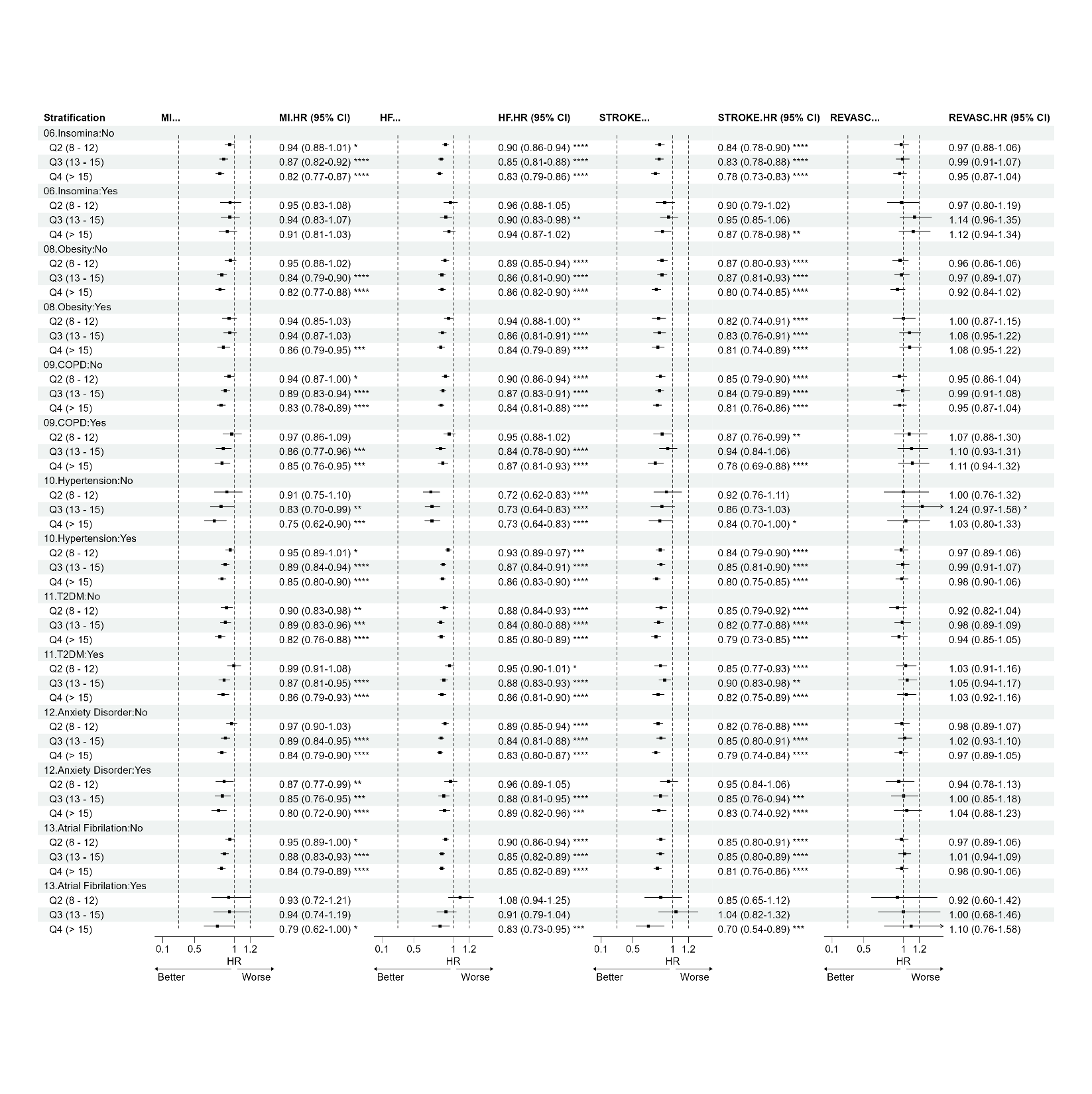

**(continued)**

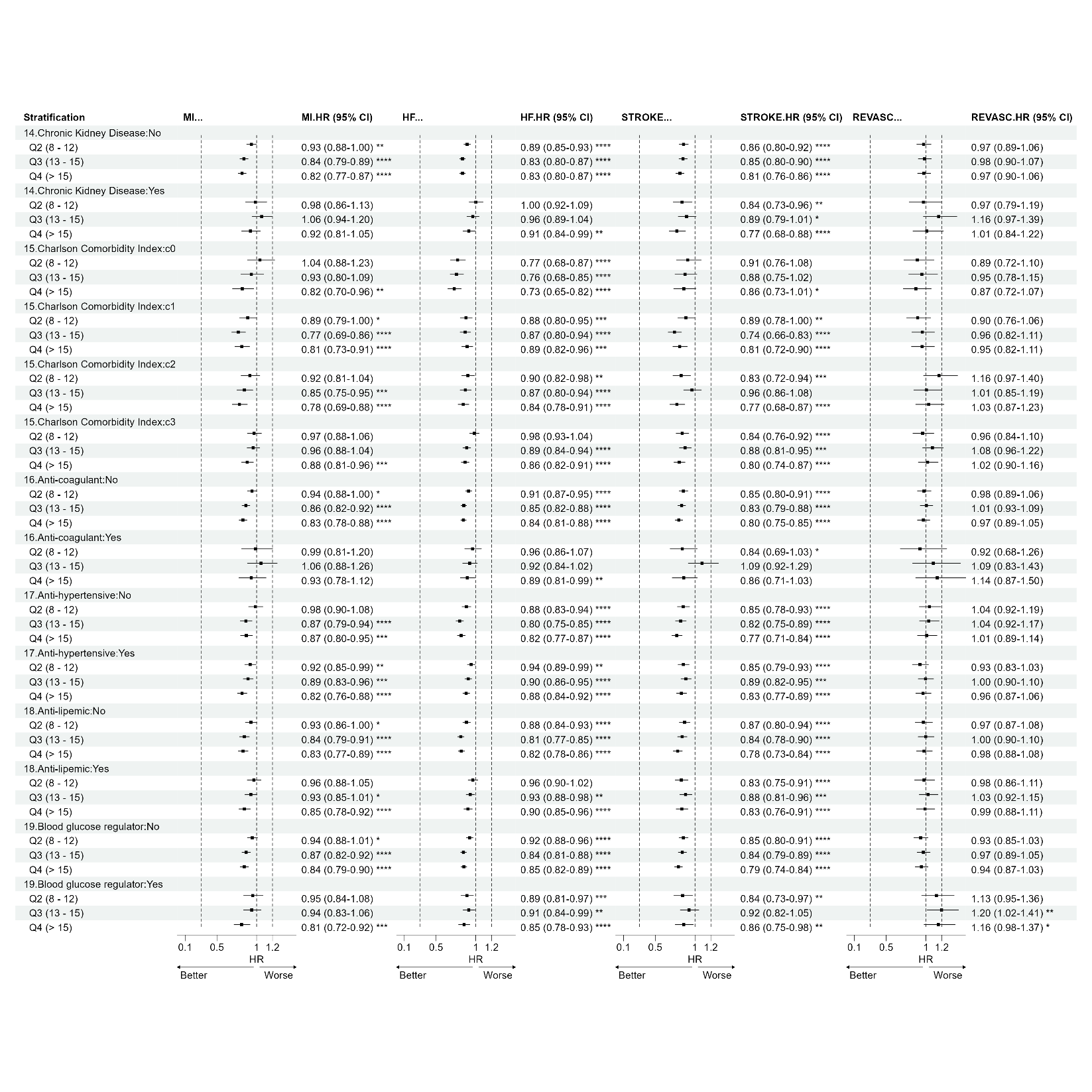

**Supplemental Figure 8:** Summary of inverse probability of treatment weights (IPTW)-adjusted Cox proportional hazards models assessing the effect of PAP exposure groups based on quartiles of PAP utilization during first year on myocardial infarction (MI), heart failure (HF), stroke (STROKE) and coronary revascularization (REVASC) within categories of relevant sociodemographic and clinical characteristics. PAP exposure groups were defined as follows: level 1 (reference): 1-7 claims; level 2: 8-12 claims; level 3: 13-15 claims; level 4: >15 claims).Results were derived from IPTW-weighted Cox proportional hazards models adjusted for age, sex, race, low-income-subsidy or dual-eligibility indicator, type 2 diabetes, hypertension, obesity, atrial fibrillation, COPD, CKD, hypersomnia, and insomnia, anxiety disorder, hypersomnia, insomnia, CCI, prescriptions of anticoagulants, antihypertensives, antilipidemic agents and blood glucose regulators. Among stratified models, stratification variable was not included as a covariate in propensity score models nor in outcome models. Reference category is no evidence of PAP initiation. Abbreviations: ACM: all-cause mortality; COPD: chronic obstructive pulmonary disease; CKD: chronic kidney disease; CCI: Charlson comorbidity index; PAP: positive airway pressure; HR: hazard ratio; CI: confidence interval.
